# A Bayesian network-based framework to uncover the causal effects of genes on complex traits based on GWAS data

**DOI:** 10.1101/2022.12.25.22283943

**Authors:** Liangying Yin, Yaning Feng, Alexandria Lau, Jinghong Qiu, Pak-Chung Sham, Hon-Cheong So

## Abstract

Deciphering the relationships between genes and complex traits could help us better understand the biological mechanisms leading to phenotypic variations and disease onset. Univariate gene-based analyses are widely used to characterize gene-phenotype relationships, but are subject to the influence of confounders. Furthermore, while some genes directly contribute to traits variations, others may exert their effects through other genes. How to quantify individual genes’ direct and indirect effects on complex traits remains an important yet challenging question.

We presented a novel framework to decipher the total and direct causal effects of individual genes using imputed gene expression data from GWAS and raw gene expression from GTEx. The study was partially motivated by the quest to differentiate “core” genes (genes with direct causal effect on the phenotype) from “peripheral” ones. Our proposed framework is based on a Bayesian network (BN) approach, which produces a directed graph showing the relationship between genes and the phenotype. The approach aims to uncover the overall causal structure, to examine the role of individual genes and quantify the direct and indirect effects by each gene.

An important advantage and novelty of the proposed framework is that it allows gene expression and disease trait(s) to be evaluated in different samples, significantly improving the flexibility and applicability of the approach. It uses IDA and jointIDA incorporating a novel p-value-based regularization approach to quantify the causal effects (including total causal effects, direct causal effects, and medication effects) of genes. The proposed approach can be extended to decipher the joint causal network of 2 or more traits, and has high specificity and precision (a.k.a., positive predictive value), making it particularly useful for selecting genes for follow-up studies.

We verified the feasibility and validity of the proposed framework by extensive simulations and applications to 52 traits in the UK Biobank (UKBB). Split-half replication and stability selection analyses were performed to demonstrate the accuracy and efficiency of our proposed method to identify causally relevant genes. The identified (direct) causal genes were found to be significantly enriched for genes highlighted in the OpenTargets database, and the enrichment was stronger than achieved by conventional univariate gene-based tests. Encouragingly, many enriched pathways were supported by the literature, and some of the enriched drugs have been tested or used to treat patients in clinical practice. Our proposed framework provides powerful a way to prioritize genes with large direct or indirect causal effects and to quantify the importance of such genes.

## Introduction

With the rapid development of genomic technologies, our knowledge of the genetic basis of complex traits has substantially improved in the past decade. Genome-wide association studies (GWAS) have proven instrumental in uncovering susceptibility genes and variants underlying complex traits ^1-3^. Traditionally, GWAS is performed in a SNP-based manner; however, as most trait-associated variants usually have minor effects and reside in the non-coding region, it has proved very challenging to translate these findings into clinical applications ^4^.

Gene-based analyses, especially those based on imputed gene expression profiles (e.g., PrediXcan), have gained increasing popularity in deciphering disease genes. However, most studies have mainly focused on univariate associations, which may be subject to the influence of confounders. Furthermore, while some susceptibility genes directly contribute to complex traits variations, others probably exert their effects through influences on other genes. This mechanistic information could provide valuable insights on biological processes and pathophysiology of traits and diseases. At present, most gene-based association studies focus on simply identifying trait-associated genes and do not go on to characterize the exact roles of these genes on phenotypes. A *‘systems’* approach that considers the contribution of not only one but a *network* of genes will facilitate our understanding of the pathophysiology of complex diseases. Such an approach would quantify the diverse direct and indirect effects of multiple genes on complex traits, provide a deep and quantitative characterization of underlying disease mechanism. The biological knowledge gained would benefit the prioritization of novel drug targets.

Recently, Boyle et al. ^5^ suggested that complex traits/diseases are ‘omnigenic’, with numerous genetic influences, some with direct (causal) effects (‘core genes’) and others with indirect effects (‘peripheral genes’) connected to the phenotype through the core ones. The model raised much attention and discussions ^6-8^. with some even questioning the usefulness of GWAS, as many genes considered as ‘susceptibility’ genes may be only tenuously connected to the disease, and such genes may be hard to differentiate from those of direct biological relevance.

In this work, we proposed a novel framework to estimate the gene-phenotype network and to quantify the direct and indirect causal effects of genes on the studied disease/trait. In addition, joint causal effects (the causal effects of each gene when intervention is performed on a group of genes) can be computed under the same framework. We leveraged GWAS-predicted expression profiles and raw expression data from a reference sample (e.g. GTEx). The framework is based on a Bayesian network (BN) approach, which produces a *directed* graph showing the causal relationships among genes and between genes and the phenotype. It aims to identify the overall causal network and quantify the effects of genes on the studied trait, including both direct and indirect effects. The inferred causal network can be used to predict the consequences of external interventions on various genes, in isolation or combination, on target traits. By pinpointing core (driver) genes from peripheral or irrelevant genes for various traits and diseases, the causal network approach can potentially predict the effects of interventions on various genes and identify novel druggable targets for various diseases.

Our proposed causal network methodology is based primarily on gene expression, since most GWAS associations are located in non-coding DNA sequences and their effects are likely to be mediated through changes in gene expression. Furthermore, gene expression data is relatively comprehensive and abundant compared to other types of omics data and, once the effects of genetic variations on gene expression have been unravelled by eQTL analyses, the gene expression changes associated with a disease can be “predicted” from GWAS data. The use of predicted gene expression is an attractive approach since gene expression studies typically have very small sample size because of the difficulties and cost in accessing the relevant tissues for the disease, and gene expression data collected after disease onset may be confounded by medication usage and other clinical/environmental factors. Our proposed approach of leveraging large-scale GWAS-predicted expression data avoids many of these issues.

We summarize our contributions below. *Firstly*, our proposed approach avoids the difficulties of building gene-disease networks from raw expression data described above, by leveraging large-scale GWAS-predicted transcriptome data. As far as we know, our study is the first to use large-scale GWAS-predicted and raw gene expression data from different samples to construct structural networks of genes and phenotypes and to estimate the strengths of the causal connection. Our proposed framework integrates data from multiple sources to infer the causal model, greatly enhancing its flexibility and applicability.

*Secondly*, the proposed method for gene-outcome causal relationship estimation (which is one component of our framework), i.e., PC-simple algorithm, has not been applied to GWAS before; it can thus be considered a novel approach to uncover important genes in the context of GWAS analysis.

*Thirdly*, previous works mainly focused on estimating structural networks between genes and phenotypes, and most did not provide quantitative estimates of the effects of genes on phenotypes. In this study, we proposed to use IDA and jointIDA incorporating a novel p-value-based regularization approach to quantify the causal effects (including total causal effects, direct causal effects, and mediation effects) of genes. The regularization approach is in fact generally applicable to any IDA/jointIDA analyses. The proposed framework has high specificity and precision (a.k.a., positive predictive value), making it particularly useful for selecting specific genes for follow-up studies.

*Fourth*, we have proposed a combination of methods to validate our framework, including split-half validation and stability selection. We also propose stability selection for statistical inference as it enables control of the expected number of false positive findings. *Fifth*, our proposed approach is general and can potentially be extended to other forms of omics data, such as proteomic or methylomic data, and to construct joint causal networks for 2 or more traits. *Sixth*, we applied the proposed framework to a wide variety of phenotypes in the UK-Biobank, and reveal the role of different genes in various diseases.

## Methods

The proposed Bayesian causal network consists of 3 parts, i.e., gene-phenotype causal network inference, gene-gene causal network inference, and penalty-based causal effects estimation(Figure1). As can be seen, we have used GTEx data for gene-gene causal network inference, and UK-Biobank data (coupled with PrediXcan and Trans-QTL) for gene-phenotype causal network inference.

**Figure 1.**
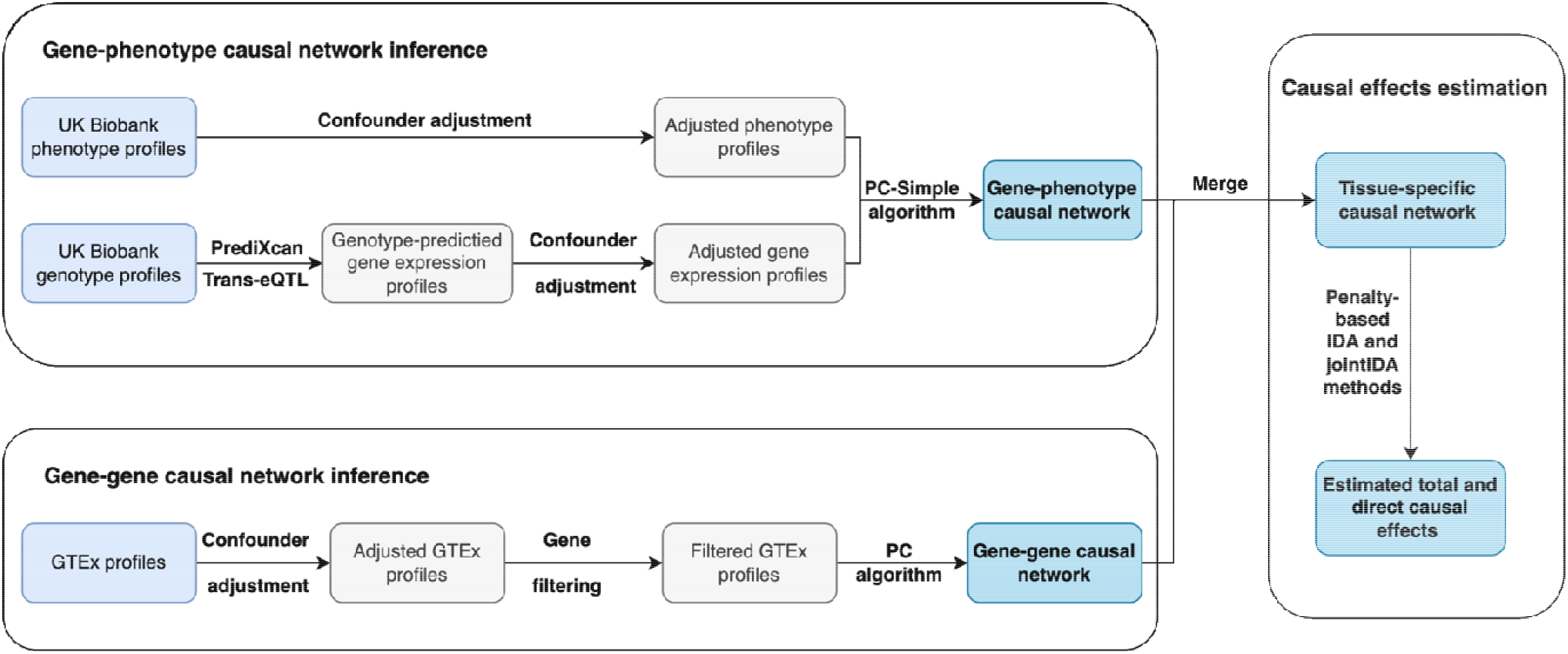
Workflow of the proposed causal inference framework based on GWAS data

Based on the assumption of independent cis- and trans-eQTL effects, We used “PrediXcan” ^9^ to map the tissue-specific cis-eQTLs, and then separately imputed tissue-specific gene expression levels from genotypic data on UK-Biobank subjects. In brief, we first built a prediction model for expression data by a regularized regression model based on a the GTEx genotype and gene expression reference data. Genotype data on UK-Biobank subjects were then inputted to the prediction model to generate and output “imputed” gene expression data on the corresponding subjects (for details, see supplementary material). Publicly available *trans*-eQTL data ^10^ was used to complement the *cis*-predicted gene expression levels. To avoid possible bias introduced by population structure, we adjusted both the genotype-predicted expression levels as well as target phenotypes by the top 10 principal components (PCs) of the corresponding genotype datasets. Then, we employed the PC-simple algorithm^11^ to infer the causal relationships between genes and corresponding traits. Briefly, PC-simple can be regarded as a generalization of correlation screening that utilizes ordered independence screening to infer the causal relationships between covariates and response. ^11^

To start with, we define a few necessary notations. Suppose Z and Y are two random variables and *S* is a variable set. Z and Y and conditionally independent if:

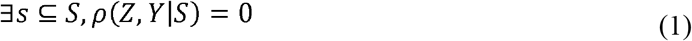

Here |*S*| defines the order of the conditional independence, *ρ* (*Z Y*, | *S*) denotes the partial correlation between Z and Y given S. Let *X* = [*X*^1^,*X*^2^, …*X*^*p*^] be a *n* × *p* matrix of adjusted gene expression data for p genes, Y be a vector of the corresponding adjusted phenotype dataset for n subjects. We could identify the gene set that are strongly associated with the target Y given other variables at a specified significance level *α* by ordered independence screening. Initially, the candidate set includes all input variables. We could iteratively update the candidate set by removing independent variables via conditional independence tests. The conditional independence tests are done level by level with increased order of the conditional set, starting with empty conditional set. As shown in Algorithm 1, the initial candidate set ***G***^**0**^ is the complete set of input variables. Candidate set ***G***^**1**^ is firstly set to ***G***^**0**^ and then updated by removing independent variables via independence test condition on the null set. Similarly, candidate set ***G***^**2**^ is firstly set to ***G***^**1**^ then updates by removing variables that are independent of Y given any other single variable in ***G***^**1**^. Through recursively performing independence screening with increased order of conditional set, we could exclude genes from previous candidate active gene set until it does not change anymore. To improve the computational efficiency of this algorithm, we set the maximum order for partial correlation screening to be 3. Fisher’s Z-transform is employed to test whether a partial correlation is zero, i.e., :

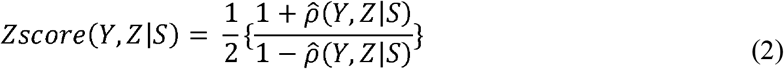

As suggested by Buhlmann et al^11^, the null hypothesis 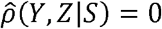 would be rejected if

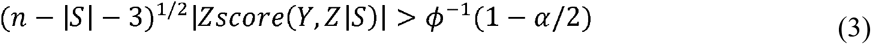

Here *α* and *ϕ* respectively denote the significance level and standard normal cumulative distribution function. In this study, we set *α* = 0.001. After employing the PC-simple algorithm, we could get the causal relationship between genes and clinical phenotypes, with a vector *Z*_*p*×1_ indicating the reliability of the inferred causal relationships. Notably, we could convert the Z score vector into a p-value vector by: *P* = 2 * *ϕ*(−*abs*(*Z*)). Please refer to the supplementary file for a more detailed description of the PC-simple algorithm.

### Algorithm 1 PC-simple algorithm

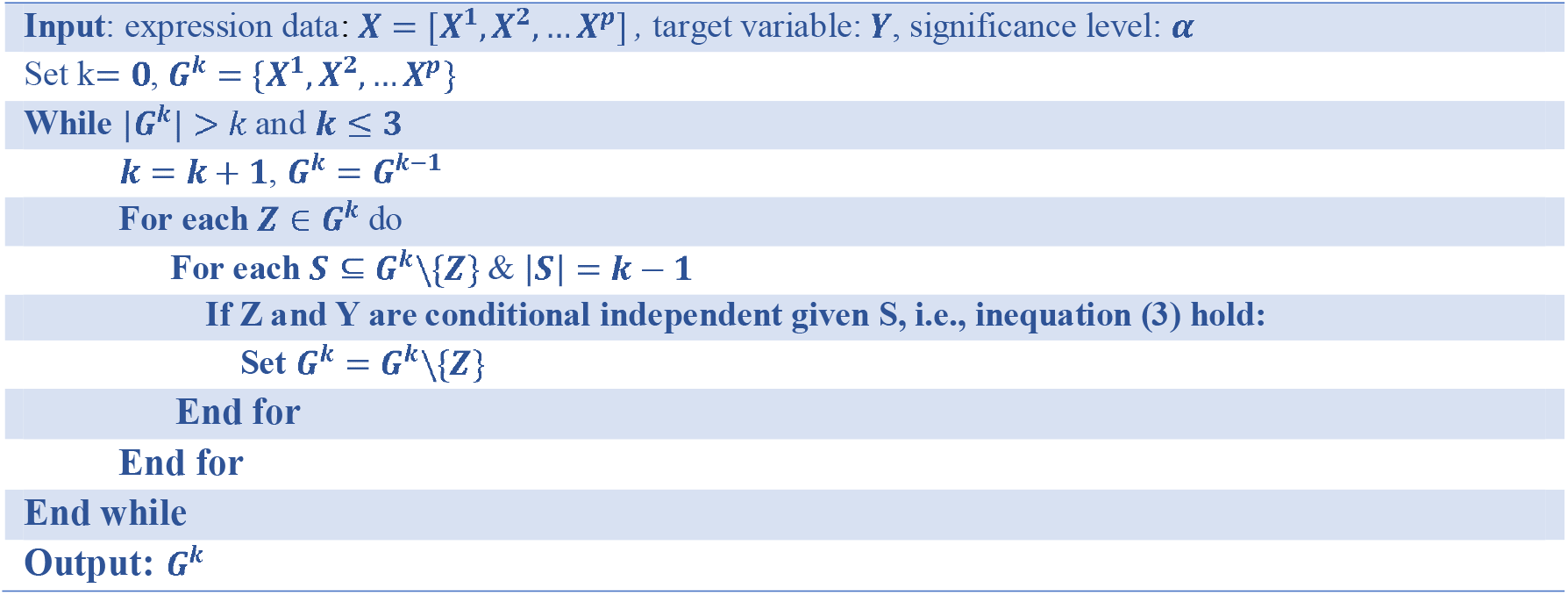

### Gene-gene causal network inference

For the gene-gene causal network inference, we used tissue-specific gene expression data from the Genotype-Tissue Expression (GTEx) database (note that any reference expression dataset can be used). To avoid possible biases introduced by confounders like population structure, batch effects, and so on, we performed confounder adjustment for the tissue-specific gene expression profiles first. This study implemented two different gene analysis strategies (exploratory and selective analysis). For the first one (exploratory analysis), we kept genes consistent with those of the gene-phenotype causal network. In other words, we took the intersection of tissue-specific genes from GTEx and the ones from imputed expression profiles. For the second strategy (selective analysis), we only selected a subset of genes from the first strategy. Suppose *N*_*c*_ is the number of identified directly causal genes for the studied traits. If *N*_*c*_ > 150, then the top 150 directly causal genes with the largest z scores will be selected. If 20 < = *N*_*c*_ < = 150, then all directly causal genes will be selected. Otherwise, the top 20 genes with the largest z scores will be selected. PC algorithm ^12,13^ will be employed to estimate the gene-gene causal graphs for the exploratory analysis while the bootstrapped-PC algorithm will be used for the selective analysis.

Graphical lasso is an efficient algorithm extensively used to estimate graphical models in high-dimensional settings. In this study, we utilized graphical lasso^14-16^ to identify independent variables in the overall gene set analysis. In brief, it was employed to enforce sparsity on our estimated causal graphs. Genes *i* and *j* are conditionally independent if the *ij*^*th*^ component of the inverse covariance matrix (∑^−1^) is zero. If we pose an L1-penalty for the estimation of ∑^−1^, we could get a sparse gene-gene network. Friedman et al.^14^ suggested setting the penalty parameter to *sprt*(log(*n*/*p*)), where *n* and *p* respectively indicate the sample size and number of variables.

Like the PC-simple algorithm, the PC-algorithm also utilizes ordered independence screening to estimate the skeleton of the underlying causal structure for high-dimensional data. To begin with, we assume that all variables were connected, i.e., we have a complete undirected graph. Then we could recursively delete edges based on the ordered independence test. After this step, we could derive a *p*-value matrix *Pval*_*p*_×*p*, which indicates the reliability of the inferred correlations between different pairs of genes. The derived undirected skeleton could be further partially directed by Pearl’s inductive causation algorithm ^17^. Precisely, for each pair of non-adjacent variables (*X*^*i*^,*X*^*j*^) with a common neighbor variable *X*^*k*^, if *X*^*k*^ does not belong to the separation set of *X*^*i*^ and *X*^*i*^, then we could orient the edges among them as:

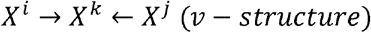

After identifying all the v-structures in the graph, we could iteratively apply Meek’s four rules ^18^ to orient as many undirected edges as possible. For more details about the algorithm, please refer to the original paper ^12,13,17,18^

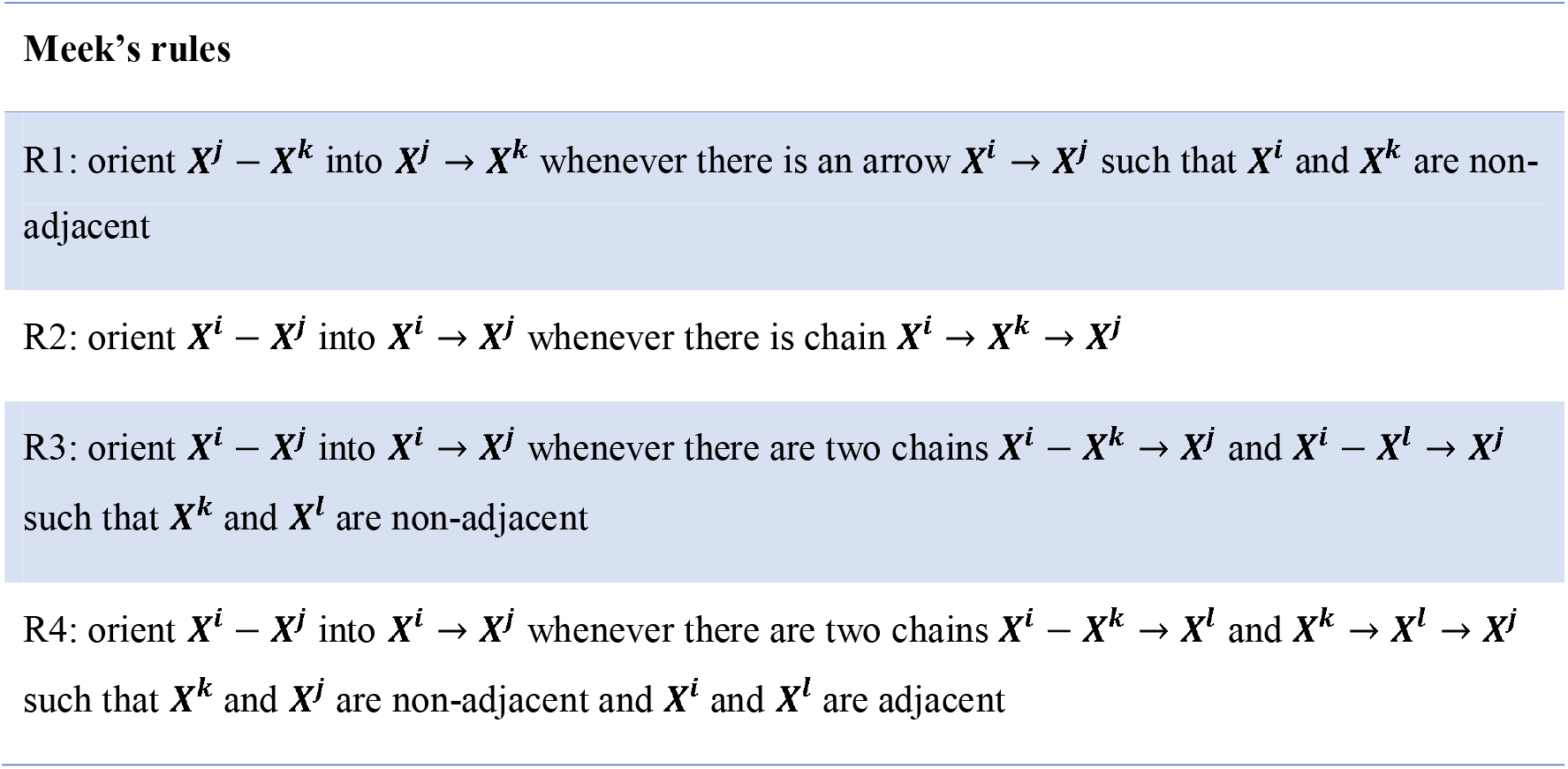

Sometimes, we may not be able to identify a valid causal graph, i.e., a completed partially directed acyclic graph (CPDAG), for the high-dimensional gene expression data. For example, there may exist circles in our derived causal graphs. We could resolve this by utilizing auxiliary methods to re-orient undirected edges to get a valid CPDAG. Also, we could incorporate domain knowledge to re-orient undirected edges. This study employed the additive noise model ^19^, which was implemented in R package “CAM”, to re-orient undirected edges. In brief, “CAM” is a structure learning algorithm that decouples order search between variables based on additive regression models. While using the “CAM” method, we selected “Lasso” as the feature selection method and linear model to estimate causal structures. Due to the comparatively small sample size, we set the number of basis functions for the pruning method to 1.

We employed a bootstrapped-PC algorithm for the selective gene analysis to infer the underlying gene-gene causal network. Briefly, we randomly resampled the available GTEx data with replacement and estimated the gene-gene network using PC algorithm. We repeated this process 100 times to get the ultimate gene-gene network. Causal relationships identified in >= 60% of the iterations were deemed actual relationships. In other words, we only retained the edge with the condition that it existed in more than 60% of the inferred causal graphs. Notably, the p-value matrix *Pval*_*p*×*p*_was the average p-value of all positive iterations. For example, if the causal relationship *i* → *j* existed in 70% of the iterations, then the ultimate p-value 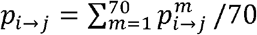, with *m* indicates the iteration index where causal relationship *i* → *j* existed.

### Penalty-based causal effects estimation

We could derive the overall gene-outcome causal network and the corresponding p-value matrix by combining the gene-gene and gene-outcome causal networks derived from the above two steps. As mentioned above, we could separately get a *p* × 1 p-value vector (*P*_*X*−*Y*_) and *p* × *p* p-value matrix (*P*_*X*−*X*_) for the inferred gene-outcome and gene-gene causal networks. The overall p-value matrix can be represented as: 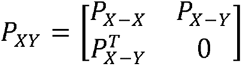. In this study, we employed the “add background knowledge” function, implemented in R package “pcalg” ^20^, to merge gene-gene and gene-outcome causal networks into one integrated causal graph.

After deriving the tissue-specific causal graph for each studied trait, we could estimate the total and direct causal effects of different genes on our studied traits. Maathuis et al. proposed efficient methods IDA ^21,22^ and jointIDA^23,24^ to infer total and direct intervention effects from high dimensional observational data based on the covariance matrix. Given a causal graph *G*, the causal effects of a particular node ***X***^***i***^ can be estimated by intervention calculus. IDA estimates the total causal effect of the intervention node ***X***^***i***^ by regressing the target on the intervention node ***X***^***i***^ and its parents node set (*Pa*(*X*^*i*^)). Given a causal graph *G*, we could find the parents node set for any given node. jointIDA is an extension of this method with multiple simultaneous interventions. When the causal graph is known, we could identify paths from node ***X***^***i***^ to the target *Y*. If we intervene on ***X***^***i***^ and all parents nodes set of the target *Y*(Pa(***X***^***i***^)) then the estimated causal effect for intervention node ***X***^***i***^ is the direct causal effect. For more details about these two methods, please refer to the supplementary text.

In this study, we proposed *novel penalty-based IDA and jointIDA approaches*, which enable us to quantify the total and direct causal effects of different genes on studied traits. More specifically, we regularized the covariance matrix by the *p*-value matrix *P*_*XY*_ derived from our inferred causal graph. Intuitively, the derived p-value matrix *P*_*XY*_ reflected the credibility of the inferred causal relationships between genes and phenotypes. The smaller the *p*-value, the more credible the estimated relationship is, and the less we penalized for the covariance. We proposed two different strategies to estimate the target covariance matrix. For the 1^st^ strategy (p-value adjustment), we obtained the target covariance matrix by solving the following optimization problem for the penalized log-likelihood of the covariance matrix:

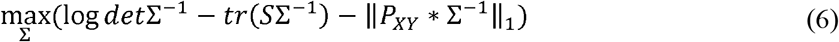

Here ∑ is the covariance matrix we aim to estimate, *S* is the estimated covariance matrix from the imputed expression profiles and raw phenotypes. * indicates component-wise multiplication. ∥*P*_*XY*_ * ∑^−1^ ∥_1_ is the L1 norm: the sum of absolute values of elements in matrix *P*_*XY*_ * ∑^−1^. For the 2^nd^ strategy (iterative p-value adjustment), we iteratively penalized the covariance matrix by the overall *p*-value matrix *P*_*XY*_ until the change of log-likelihood of the covariance matrix was less than 10^−1^, i.e.,

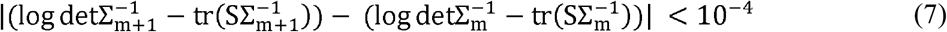

Here *m* = 1,2,3, … denotes the number of iterations. After deriving the penalized covariance matrix, we could respectively estimate the total and direct causal effects of various genes on the studied traits by IDA and jointIDA.

### Simulation study

To verify the feasibility and validity of our proposed framework, we simulated different scenarios of causal networks with various genotypes (single nucleotide polymorphisms (SNPs), genes), causal effects, graph densities, and available sample sizes.

#### Causal structure simulation

In our study, a graphical model was used to represent the causal relations between genes and phenotypes. Firstly, we randomly generated a directed acyclic graph (DAG), in which the nodes represent the variables that we are studying (including genotypes, genes, and phenotype), and the weight refers to the causal effect. All the causal effects were randomly generated by a given range (minimum and maximum weight), following the uniform distribution. We fitted 20 replicates to all combinations of:

Number of SNPs ∈ {500,700,1000,1500}

Gene number ∈ {50,70,100,150}

GTEx sample size ∈ {800, 2000, 8000}

Graph density ∈ {0.02, 0.05, 0.1}

Minimum causal effect size ∈ {0.1, 0.3, 0.5}

Maximum causal effect size ∈ {0.1, 0.3, 0.5}*1.2

Table S1 lists all the scenarios that we simulated.

#### Strategy for sample generation

Simulated samples were generated for further analysis based on the causal structure of DAG. Multivariate datasets with dependency structures were generated by the function ‘rmvDAG’ from R package ‘pcalg’ based on the earlier parameter settings. For each scenario, we divided the simulated dataset into two subsets, A and B. The sample size of dataset A was set to 800, which is close to the available sample size from Genotype-Tissue Expression (GTEx) database. We imputed gene expression data by training a prediction model in dataset A, and applied it to the genotypes of dataset B. The predicted gene expression dataset was marked as dataset C. An elastic-net regularized regression model was used for the prediction model. Notably, cross-validation was utilized to tune the optimal parameters for the prediction model. These two datasets, A and C, were separately used for further gene-gene and gene-phenotype causal network inference. R package ‘glmnet’^25^ was used in this section. Different scenarios were tried with varied GTEx sample sizes (More details are provided in Table S1).

#### Causal structure inference

We applied our framework to the simulated datasets to estimate the causal structure. PC-algorithm was used to infer the gene-gene network based on actual expression data (dataset A). The PC-Simple algorithm was used to infer the gene-phenotype network based on imputed expression data (dataset C). Then we merged the inferred gene-gene and gene-phenotype networks to get the overall causal structure. After obtaining the overall causal structure, total and direct causal effects could be estimated for further analysis.

#### Framework performance evaluation

Univariate tests were also performed on the imputed expression data to reveal trait-associated genes. They were widely used for gene-based analyses to identify trait-associated genes. Then, we performed different comparative analyses between univariate test and our proposed framework, including causal effect estimation and core gene detection. We calculated the correlations and RMSEs between the actual and estimated effects from both methods to compare their performances.

Notably, we applied the proposed regularization strategies (p-value matrix and iterative p-value adjustment) to estimate the covariance matrices (please refer to ‘Penalty-based causal effects estimation’ for more details). The estimated covariance matrices were used to calculate the total and direct causal effects.

As for core gene detection, we compared the differences between our framework and the univariate test in terms of sensitivity, Type I error, positive predictive value (PPV), negative predictive value (NPV).

Also, we are interested in finding mediating genes, which could provide valuable information for understanding the biological mechanisms of complex traits. Therefore, mediator detection ability was also evaluated. Again, sensitivity, Type I error, PPV and NPV were separately calculated to demonstrate the performance of our proposed method in uncovering mediating genes.

### Split-half replication and stability selection analysis

Split-half replication was performed to assess the robustness and validity of our method in uncovering causally relevant genes for studied phenotypes. Firstly, we randomly split the dataset for our studied phenotypes into 2 subsets. Then our proposed framework was applied to these two subsets independently. We evaluated the validity and robustness of our proposed framework by calculating the replication rate of the identified directly causal genes from these two subsets for the same traits.

Apart from this, stability selection ^26^ was performed to quantify the stability of our proposed method in identifying core genes or causal relationships. This approach also provides statistical inference as it can control the expected number of false positives (i.e. E(*V*)). In brief, it combines subsampling with a high-dimensional feature selection algorithm to improve the accuracy of the estimated causal structure and control for the expected number of false positive findings. Notably, core genes could also be regarded as causal relationships between genes and clinical outcomes. Suppose there are *p* variables and *n* samples for our structure estimation problem. *I*_*b*_ indicates a random subset of the samples with size *m*= ⌊ *n*/ ⌋. 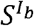 denotes the set of identified core genes/causal relationships by our proposed method based on subset 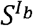. The set of all identified core genes/causal relationships can be expressed as: 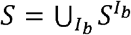. Let *q* be the average number of identified core genes/causal relationships, 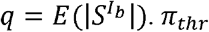 defines the cutoff that is used to determine whether an inferred core gene (causal relationship) is falsely identified. The number of falsely identified core genes/causal relationships E(*V*) under *π*_*thr*_ is bounded by:

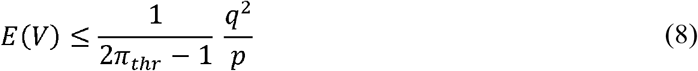

As suggested by Meinshausen et al. ^26^, *π*_*thr*_ has an almost negligible effect on the results when it falls into the sensible value range [0.6, 0.9]. In this study, the cutoff (*π*_*thr*_) was set to 0.6. By employing this method, we could quantify the average number of identified variables (*q*), the number of false identified variables(*E(V)*), and the expected proportion of false positives when compared to the (average) number of selected features, *E*(*V*)/*q*.

### OpenTargets enrichment analysis

To further evaluate the reliability and accuracy of our proposed method in identifying causally relevant genes, we performed enrichment analysis of potential targets associated with studied disorders. The targets of corresponding disorders were downloaded from the Open Targets Platform. Open targets provide us with measures of relevance (from 0 to 1) between diseases and targets based on various factors, like associations with known drugs, hits from GWAS (genome-wide association study), RNA expression studies, etc. Specifically, we tested whether the identified directly causal genes from our proposed framework were more likely to be targeted for the corresponding disorder. In addition, we compared the identified directly causal genes from our framework with the susceptible genes identified from the univariate test. Since our proposed method is more stringent regarding defining trait-associated genes, the number of identified genes from our method tends to be much smaller than that from the univariate test under the same p-value cutoff. We selected the same number of genes for comparison, to avoid possible bias due to imbalanced sample sizes. Concretely, we ranked the analysis results from the univariate test by p-value in an increasing order first. Then we select the top *N*_*C*_ genes, where *N*_*C*_ equals the number of identified directly causal genes from our proposed framework.

### Survival and replication analysis for breast cancer

Apart from the above-mentioned enrichment analysis, we also performed a survival analysis for breast cancer to verify the validity of our proposed framework. Cox proportional hazard model was used to estimate the influence of the identified causally relevant genes from our proposed framework on the survival of cancer patients. We used experimental RNA expression levels from the Kaplan Meier Plotter database as the source to validate the physiological importance of the identified genes from our framework. There are 4929 available patients for the survival analysis, which is much larger than The Cancer Genome Atlas (TCGA) dataset. In this study, the survival analysis was performed using the default settings. Additionally, we performed survival analysis on gene sets identified from the univariate test. Similar to open target enrichment analysis, the same number of genes were separately selected for breast mammary and whole blood tissues based on derived p-values. Then we compared the survival analysis results derived from our method and that from the univariate test. Furthermore, we examined whether our proposed method could identify more pathogenic genes than expected by the Fisher’s exact test. To be more precise, we compared the identified core genes from our method with the known pathogenic genes database (OncoVar ^27^) for breast cancer.

### Further analyses

Our proposed method could identify both directly, indirectly causal genes and quantify their effects on the studied traits. To better understand the underlying biological mechanism of the studied traits, we conducted pathway enrichment analysis on the identified gene set by the “ConsensusPathDB”^28^ platform. More specifically, over-representation analysis was performed on the directly causal gene set by hypergeometric test. In addition, enrichment analyses were performed by “Enrichr” ^29,30^ to identify transcription factors and drugs related to studied binary traits. As we mentioned before, gene-based analyses, especially those based on imputed expression profiles, are mainly based on the univariate test. In this study, we performed univariate analyses for all studied tissue-trait pairs. Then we compared the susceptible gene set identified from the univariate test with that from our proposed method. Also, we compared the estimated effects of these genes on the studied traits. Notably, our proposed framework could incorporate external knowledge to estimate the causal graph. In this study, we performed exploratory analyses on whole blood-related traits by incorporating external knowledge from SIGNOR ^31^.

### Multiple traits analysis

The proposed causal inference framework could also be extended to more than one trait. Here, we take two traits as the demonstration example. Firstly, we need to identify the causal directions between the studied traits, which could be inferred from Mendelian Randomization or extracted from literature. Then, we picked out the outcome trait as the “ultimate” outcome. As for the exposure trait, we first calculated the polygenic risk scores (PRS) of all involved subjects based on external GWAS summary data. Then we randomly selected 5,000 subjects to build a prediction model to predict the exposure trait based on calculated PRSs. Subsequently, we applied the built model to predict the value of exposure trait for the remaining subjects. In this study, a linear model was used to predict the value of the continuous exposure trait, while a logistic model was used for the binary exposure trait. The predicted exposure trait could be regarded as a special (imputed) ‘gene’. After that, we combined the predicted exposure trait with the imputed gene expression profiles to infer the “gene”-outcome causal network. The whole graph for the exposure trait could be regarded as the special “gene-gene” causal network. The overall causal graph could be derived by combining the inferred “gene”-outcome network and “gene-gene” network. The workflow for the causal graph inference for more than 2 traits is like that for 2 traits. Similar to single-trait analysis, we could quantify the total and direct causal effects of various genes on the “outcome” trait.

## Results

### Simulation results

#### Results of causal effects estimation

Firstly, we compared the correlation and root mean square error (RMSE) between the actual and estimated value. Fig.2 shows the comparison results of total causal effect (IDA) and direct causal effects (JointIDA) for 20 replicates. As shown in Fig.2, our proposed p-value regularization strategy could significantly improve the accuracy of total causal effects estimation. More specifically, for the GTEx sample size =800, the median correlation of 20 replicates increased from 0.7 to 0.82, while the RMSE decreased from 1.26 to 0.68. Correlation and RMSE improved with increased GTEx sample size. A similar trend was observed for the estimation of direct causal effects (Fig.2C and D). Obviously, our proposed framework performed well in quantifying the total and direct causal effects of various genes on the studied trait. Most importantly, it consistently achieved superior performance than the univariate analysis.

**Fig. 2.**
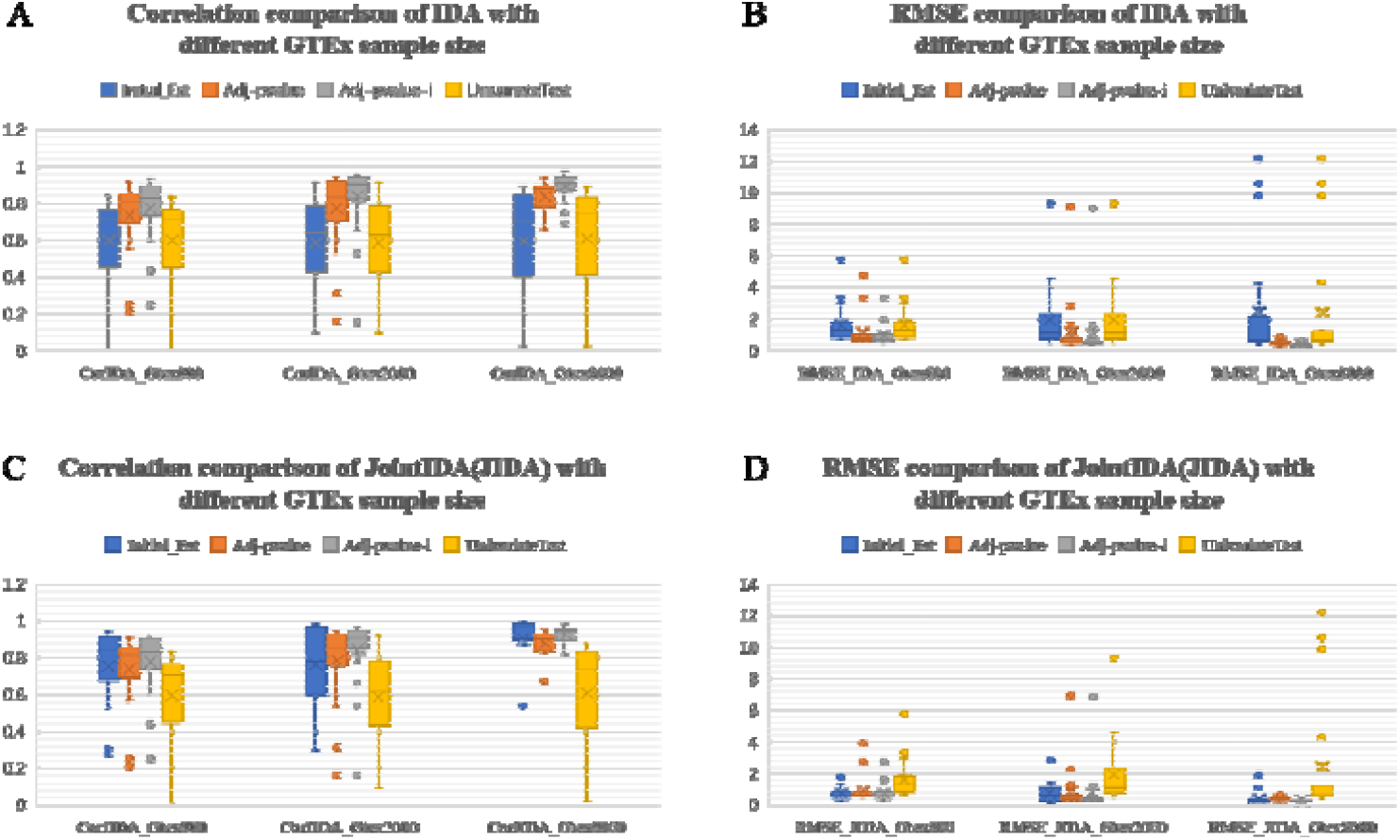
The correlation and root mean square error (RMSE) of IDA and JointIDA (JIDA) comparison between true and estimated value with different GTEx sample number. (1) A: correlation between true IDA and estimated IDA. B: RMSE between true IDA and estimated IDA. C: correlation between true JointIDA(JIDA) and estimated JointIDA(JIDA). D: RMSE between true JointIDA(JIDA) and estimated JointIDA(JIDA). (2) Four methods were employed to obtain the estimated value. ‘Initial_Est’ refers to the estimated value without any adjustment. ‘Adj-pvalue’ refers to the estimated value after adjusted by the p-value matrix. ‘Adj-pvalue-i’ refers to estimated value after iteratively adjusted by the p-value matrix. ‘UnivariateTest’ refers to the estimated value from univariate analysis. (3) We obtain the estimated value based on different GTEx sample size. ‘GTEx800’ refer to the scenarios with 800 samples for Gene-Gene causal network inference, while ‘GTEx2000’ refer to the scenario with 2000 samples. And so on. (4) Parameter setting: genotype number is equal to 700, gene number equal to 70, overall sample size is 100000, graph density is equal to 0.05, min weight in graph is 0.1, max weight is equal to min weight*1.2. The boxplots were generated based on 20 replicates.

The p-value adjustment strategy significantly improved the accuracy of estimated causal effects for genes from IDA and JointIDA methods, especially those without any causal effects in the actual graph. Nevertheless, in some special cases, some genes have both direct and indirect causal effects on phenotypes (or one gene has causal effects on phenotype according to several mediating genes). These causal relations cannot always be accurately inferred in the estimated graph because of small sample sizes. In these cases, the p-value adjustment strategy may have limited power in improving the estimation accuracy, resulting in some outliers in the boxplot. Based on the simulation results, this could be alleviated by increasing the available GTEx sample size. For more details, please refer to Table S2-3.

Fig.3 shows the comparison results with varied gene numbers. The performance of our framework was relatively stable as the gene number increased. Generally, the actual and estimated IDA correlations were around 0.75 after applying the p-value adjustment strategy. We also compared the results from different perspectives, including varying graph densities, causal effect sizes (Fig.S1-11). Overall, our proposed framework performed well in uncovering the causal effects of genes on studied traits under various parameter settings.

**Fig. 3.**
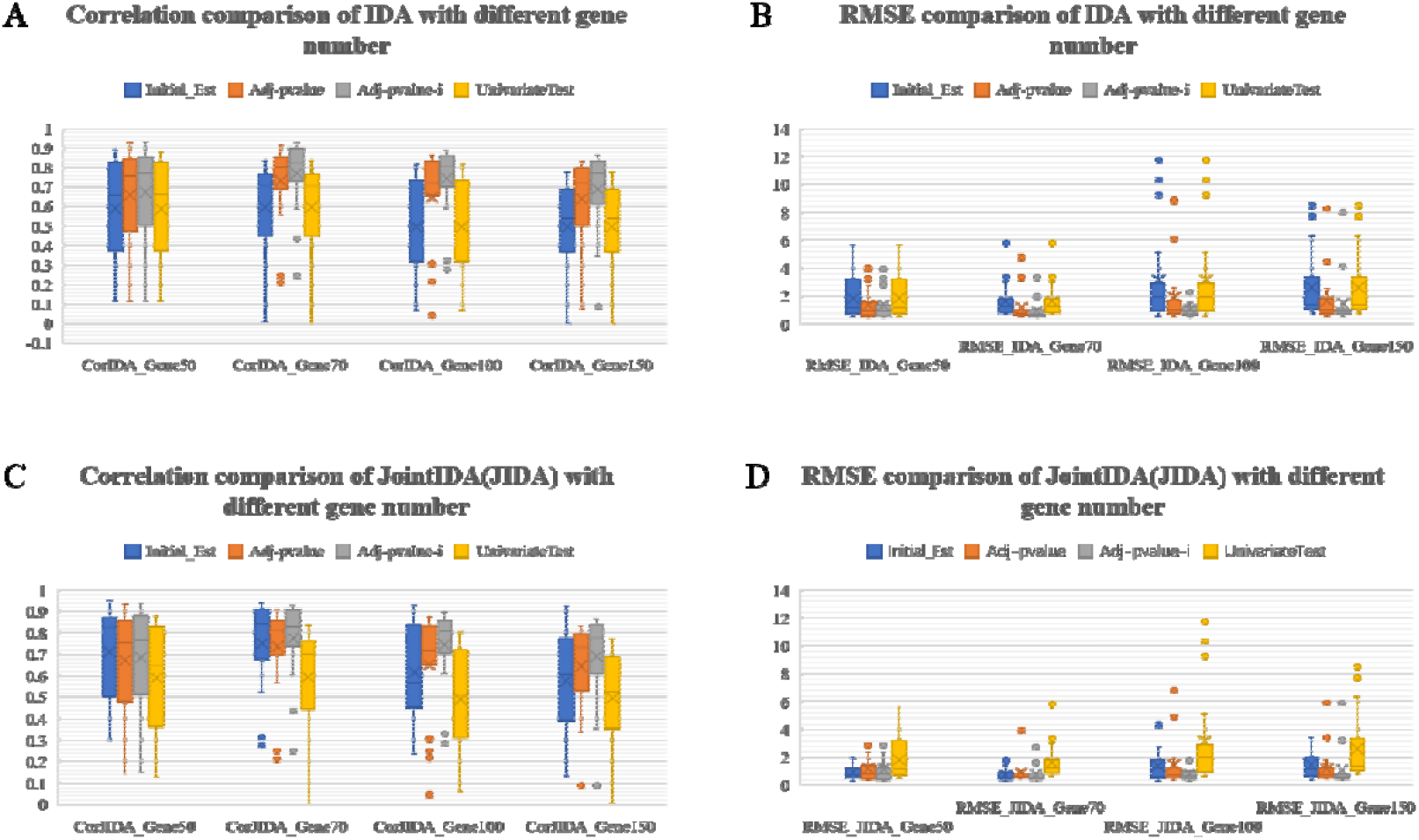
The correlation and root mean square error (RMSE) of IDA and JointIDA (JIDA) comparison between true and estimated value with different gene number. (1) A: correlation between true IDA and estimated IDA. B: RMSE between true IDA and estimated IDA. C: correlation between true JointIDA(JIDA) and estimated JointIDA(JIDA). D: RMSE between true JointIDA(JIDA) and estimated JointIDA(JIDA). (2) Four methods were employed to obtain the estimated value. ‘Initial_Est’ refers to the estimated value without any adjustment. ‘Adj-pvalue’ refers to the estimated value after adjusted by the p-value matrix. ‘Adj-pvalue-i’ refers to estimated value after iteratively adjusted by the p-value matrix. ‘UnivariateTest’ refers to the estimated value from univariate analysis. (3) We obtain the estimated value based on different gene number. ‘Gene50’ refers to the gene number is equal to 50, and so on. (4) Parameter setting: overall sample size is 100000, GTEx sample size is 800, graph density is equal to 0.05, min weight in graph is 0.1, max weight is equal to min weight*1.2. The boxplots were generated based on 20 replicates.

#### Results of core genes detection

Accurate identification of directly causal genes to studied phenotype is essential for further study. Our framework used imputed gene expression data (instead of raw gene expression data, because of the limited sample size) to infer the gene-phenotype network. We also conducted comparison analysis for the PC-Simple and univariate test.

Fig.4 shows the comparison results between the PC-Simple and univariate test in identifying trait-associated (or direct causal) genes. As shown in Fig.4, PC-Simple had superior performance than the univariate test in uncovering relevant genes. It had a much higher positive predictive value (PPV) and a lower false positive rate (FPR). The performance improved with an increased GTEx sample size.

**Fig. 4.**
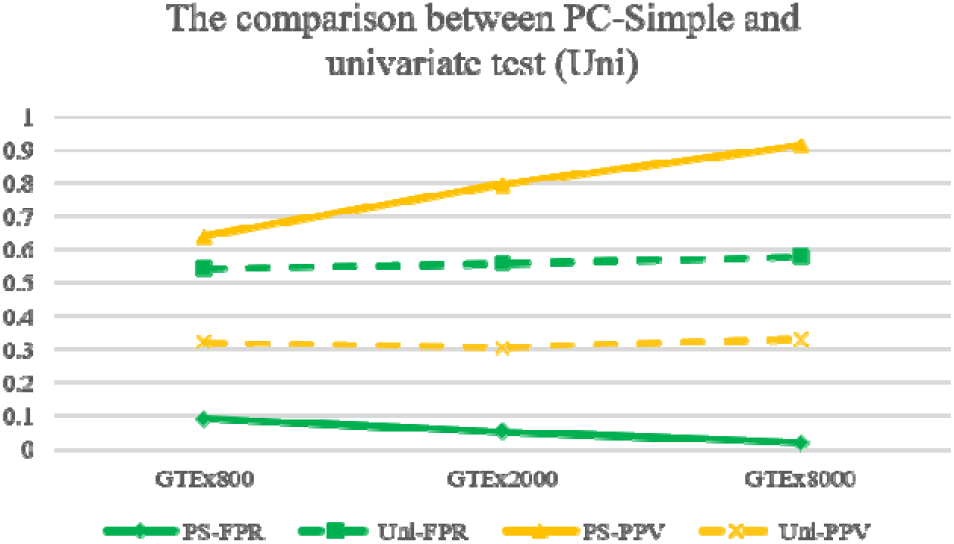
The comparison between PC-simple and univariate test. *FPR: false positive rate; PPV: positive predictive value. Parameter setting info: genotypes number is equal to 700, gene number equal to 70, overall sample size is 100000, graph density is equal to 0.05, min weight in graph is 0.1, max weight is equal to min weight*1.2. The boxplots were generated based on 20 replicates.

Table 1 demonstrates the performance of the PC-Simple and univariate test with different gene numbers. Obviously, the performance of the PC-Simple remained stable with increased gene numbers. Even though the sensitivities for the PC-Simple method were in general lower than the univariate test, it achieved high accuracy. Since the PC-Simple method was more stringent than the univariate test regarding the definition of associated genes, it tended to select fewer genes, leading to lower sensitivity. Most importantly, the type I errors for the PC-Simple method were all significantly lower than the univariate test.

#### Results of mediator detection

We simulated different scenarios to demonstrate the performance of our proposed method in mediator detection. Table 2 shows the results of mediator detection with varied gene numbers (50,70,100,150), graph densities (0.02, 0.05, 0.1), causal effect sizes (0.1, 0.3, 0.5), and GTEx sample sizes (800, 2000, 8000). Overall, our proposed method performed well in detecting mediators. The PPV for mediator detection of our framework can reach 0.88, while the Type1 errors were controlled around/under 0.1. The sensitivity and PPV were relatively stable with increased gene numbers. In view of our computational power limit, the max gene number in all simulated scenarios was set to 150. Since the performance of our framework is not sensitive to gene numbers, we believe it will still have satisfactory performance with even larger gene numbers.

#### Results of further analyses

The accuracy of the estimated graph could be affected by the available GTEx sample size. To improve the performance of our framework, we used bootstrap for the inference of gene-gene networks. Precisely, we resampled our simulated dataset with replacement and estimated the gene-gene network. We repeated this process 100 times to get our final gene-gene network. As mentioned earlier, we only retained edges that can be learned from more than 60% of the estimated graphs. Table 3 shows the performance of our proposed method in identifying gene-gene interactions with and without bootstrap. We compared the sensitivity, Apparently, bootstrap could significantly improve the accuracy of identified causal relationships between genes, with median PPV increased from 0.57 to 0.9.

Table 4 demonstrates the results of mediator detection under two different strategies. Again, bootstrap significantly improved the performance of the proposed method in detecting mediators. To be more specific, median PPV increased from 0.7 to 0.935 while the Type1 error decreased from 0.34 to 0.038.

### Application to UKBB phenotypes

We applied our proposed approach to 41 continuous and 11 binary traits. Details about the studied phenotypes are summarized in Table 5. The individual-level phenotypes and genotypes data were obtained from the UK Biobank (UKBB) with application no. 37268. In this study, continuous traits were directly extracted from the UKBB dataset. The binary traits diabetes (UDI:2443-0.0) and COVID-19 were also directly extracted from the UKBB. Notably, for COVID-19, we considered two different analysis strategies. For the first analysis strategy, cases were defined by the presence of COVID-19, while controls were subjects without COVID-19. For the second one, cases were defined by hospitalized/severe COVID-19 subjects, while controls were defined by non-hospitalized COVID-19 subjects and subjects without COVID-19. The remaining binary traits were defined by a combination of self-reported and ICD10-coded diseases. For more details about the combination of self-reported and ICD-10 coded diseases, please refer to Table S4. Notably, the case sizes of all 11 binary traits were larger than 2,000.

The inferred causal graphs were tissue specific. In this study, we imputed the gene expression levels of UKBB subjects based on genotypes for several different tissues. Concretely, we estimated the gene expression levels of UKBB subjects in the whole blood, adipose subcutaneous, adipose visceral omentum, liver, lung, pancreas, breast mammary, prostate, cortex, frontal cortex, artery coronary, heart left ventricle, nucleus accumbens basal ganglia, putamen basal ganglia. The gene expression levels for these tissues were imputed because they have been reported to be relevant to our studied phenotypes. We inferred the causal graphs for all 52 phenotypes for the whole blood tissue. As for the remaining tissues, we only inferred the causal graphs of traits that have been reported to be relevant to the corresponding trait.

Table 6 and Fig. 5 summarize the identified causally relevant genes for our studied phenotypes under the exploratory analysis. As shown in Fig. 5, some traits, especially these anthropometric-related traits, tended to have more causally relevant genes than other traits regardless of tissue types. For example, the number of identified causal genes for weight and BMI were top ranked in all relevant tissues. On the contrary, simple lab test traits, like Oestradiol and lipoprotein A, were more likely to have less causally relevant genes. Overall, the number of identified directly causal genes for all traits except one was less than 150.

**Fig. 5.**
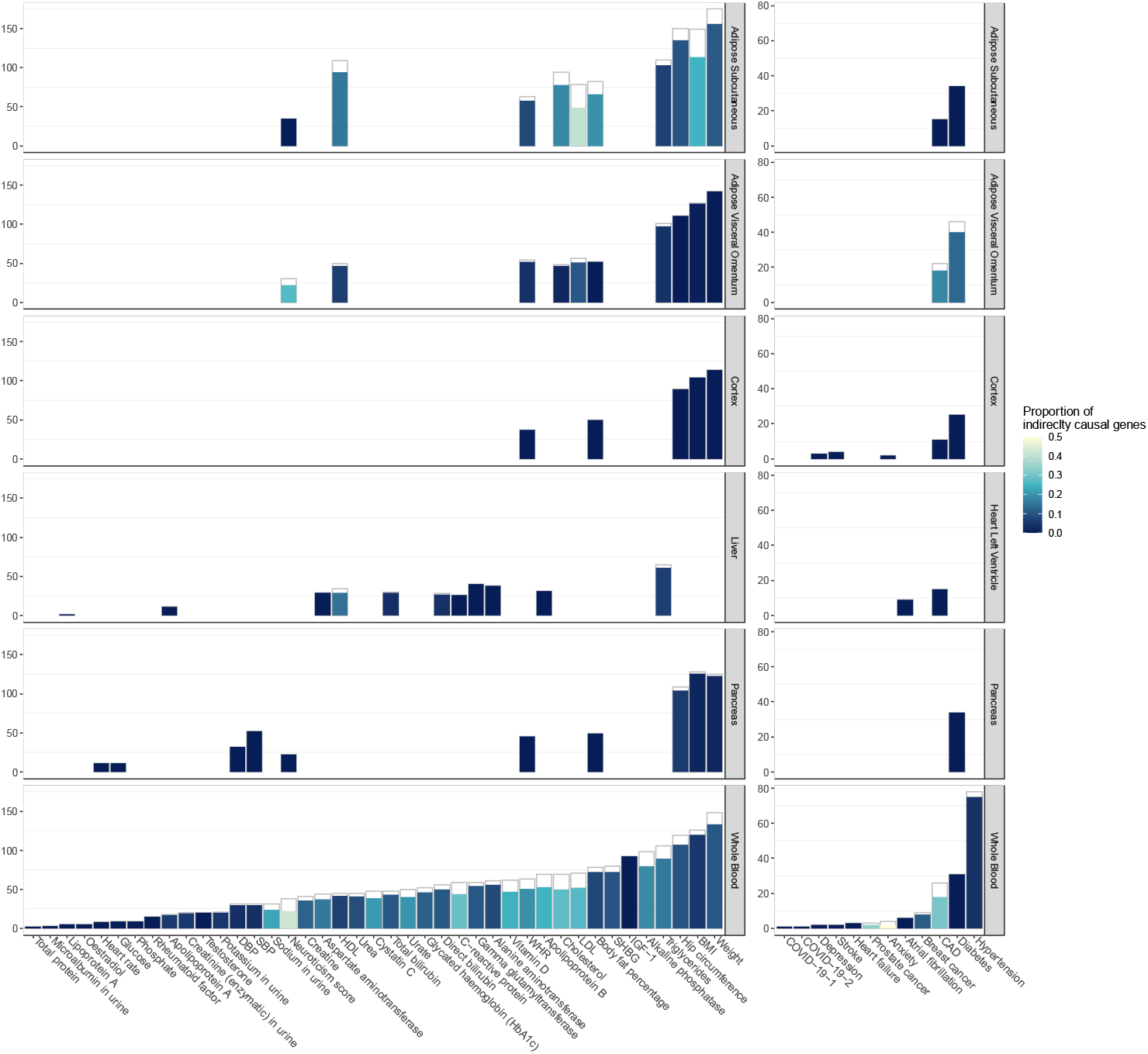
Number and proportion of identified causally relevant genes for various traits in different tissues. The length of the inner bars scale with the number of identified directly causal genes. The length of the outer bars scale with the total number of identified causal genes. The lighter the color, the higher the proportion of indirectly causal genes.

The numbers of identified causally relevant genes for binary traits were in general less than that for continuous traits. This was expected due to the much smaller effective sample size for these studied binary traits. In the UKBB dataset, the sample sizes for continuous traits were, in general, no less than 400,000. However, most of the subjects in the dataset were controls for binary traits. The cases sizes were generally less than 20,000, most of them were even smaller than 10,000. Among the 11 binary traits, only 4 of them had case sizes larger than 20,000. As can be seen from Table 5 and 6, the power of our proposed method improves with the increase of effective sample sizes.

Some traits were more likely to have a complex causal graph. In other words, they tended to have a high proportion of indirect causal genes. Usually, they were well-accepted complex traits, e.g., the proportion of indirect causal genes for neuroticism score in whole blood tissue is 42.1%. Things were different for comparably simple traits. The proportion of indirectly causal genes for these traits tended to be small or negligible. Take IGF-1 for example, we didn’t find any indirect causal genes in the inferred causal graph for the whole blood tissue. Notably, causal graphs inferred from the whole blood and adipose usually had a high proportion of indirect causal genes compared with other tissues. We assume that the complexity of the inferred tissue-specific gene-gene network may also be affected by the available sample sizes in the GTEx dataset. Whole blood and adipose are the top ranked tissues regarding the available sample size in the GTEx. Thus, the inferred gene-gene networks for these tissues had a better chance to reflect the actual interactions between various genes.

Most of the identified causally relevant genes for each trait only have a single action path to the related trait (e.g., LDL in the whole blood shown in Fig S12). At the same time, there also existed a few genes which could regulate the related trait through multiple paths. In other words, some identified genes were both directly and indirectly causal. Please refer to the following site for more examples of the inferred causal graphs: https://github.com/LiangyingYin/BayesianNetwork/tree/main/Causalgraphs.

The identified causally relevant genes for the same trait may vary between tissues. While some causal relevant genes were shared among different tissues, others were specific to unique tissues. Take BMI for example, we separately identified 114 and 120 directly causal genes for BMI in the adipose subcutaneous and whole blood tissues. Among these identified causally relevant genes, 20 of them were shared between two tissues. These shared genes were significantly enriched in the centriole, which has been proven to be involved in regulating body size^32^.

The estimated total and direct causal effects for various genes on the studied traits for the exploratory analysis were summarized in Table S5. Since we are primarily interested in the contribution of different genes to traits variations, we only estimated the causal effects of genes connected to the studied traits. While some of the identified core genes had substantial effects on studied traits, others only had minor/mild effects. For example, ALDH2 was found to have strong effect on CAD in the whole blood, while IL20RB only had minor effect. In this study, we regarded genes whose estimated total effect on the studied trait was smaller than one-tenth of the largest estimated effect as small-effect genes. Some traits were more likely to have a high proportion of small-effect core genes while others were not. Again, they usually were well-accepted complex traits. For example, the proportions of small-effect core genes for WHR (waist-hip ratio) in all relevant tissues were around/over 60%. For more details about the proportion of the small-effect genes for all studied traits, please refer to Table S6. Also, we noticed that the proportion of small-effect cores genes was high for binary traits, usually no less than 45%. The proportion of breast cancer in whole blood even reached 75%. Compared with cardiometabolic disorders, the estimated effect sizes of core genes for psychiatric disorders were much smaller. For example, the maximum absolute total causal effect of the core genes for depression was only one-fifth of that for CAD in the whole blood. Notably, the estimated causal effects for shared core genes for the same trait may vary between tissues. However, most of them usually have the same direction of effects. As expected, the estimated direct causal effects for “peripheral” genes were, in general zero, or very close to zero. However, some of them had noticeable total causal effects on our studied traits. While most peripheral genes had almost negligible causal effects, some still had relatively strong mediating effects. With the estimated total and direct causal effects of the trait-associated genes, we could have a better understanding of the contribution of these genes to the development of a particular disease (or trait variation). Most importantly, these estimated causal effects could be utilized to prioritize genes as targets for new drugs and therapeutic treatment.

Table S7 summarized the estimated causal effects of various genes on studied traits for the selective analysis. As the numbers of identified directly causal genes for all traits excluding weight in adipose subcutaneous were less than 150, the selective analysis included all directly causal genes for almost all studied traits. Compared with exploratory analysis, the proportion of both directly and indirectly causal genes were much higher for most traits. In other words, many genes, which only directly affected the studied traits in exploratory analysis, were now perceived to exert their effect on the studied trait through multiple paths. This is expected due to the confounding effects of the missing genes. Even so, the estimated total causal effects for most genes were close to that estimated from an exploratory analysis.

#### Results of split-half replication and stability selection analysis

In this study, we performed a split-half replication analysis on 9 different traits in the whole blood tissue. Table 7 summarizes the replication result. As expected, the replication rate varied between different traits. The replicate rates for continuous traits were in general high. For example, the replication rate for HDL was around 70%. Contrary to this, the replication rate for binary traits was comparatively lower. This is expected due to the more complex nature of these binary traits. Many genes may exert their effects on the risk factors of these binary traits instead of directly affecting them. It has been reported that about 30% of the identified susceptible genes for CAD were found to be involved in the metabolism of its risk factors like low-density lipoproteins (LDL), triglyceride-rich lipoproteins (TRL), and so on ^33^. On the other hand, some core genes may only have small/moderate effects on the studied traits, which made it difficult to replicate these identified core genes from a different dataset. While a small set of large effect genes stand out all the time, different sets of small/moderate effect genes may contribute to trait variations in different datasets. Apart from this, inadequate statistical power may also lead to unfavorable replication rates for these binary traits. Compared with continuous traits, the effective sample sizes for binary traits are tiny. This situation becomes even worse after we split the corresponding dataset into 2 different subsets. Notably, the replication rate for neuroticism score was also very low. Again, this may be due to the complex nature of this trait. In fact, the proportion of indirectly causal genes for neuroticism in the whole blood tissue was pretty high (42.1%). Overall speaking, the replication rates for most continuous traits were rather good. Most importantly, the proportions of shared genes for all traits were significantly higher than expected. This analysis further corroborated the effectiveness and robustness of our proposed method in identifying causal relevant genes for different phenotypes.

Stability selection was performed to quantify the stability of our proposed method in identifying core genes, with the iteration number for subsampling set to 50. Table 8 summarizes the stability selection results for 4 different traits in the whole blood tissue. As expected, the number of falsely identified core genes increases with the average number of identified core genes. Overall, our proposed framework performed well in identifying core genes. The type I errors were kept under 0.05 for all 4 traits. Apart from this, we performed stability selection to evaluate the reliability of our proposed method in identifying causal relationships between genes for selected gene set analysis. The iteration number for subsampling was set to 100. Again, our method performed well in identifying causal relationships between genes, the type I errors were all kept under 0.05 for all studied traits (Table S8). Notably, we did not assess the robustness of our proposed method in identifying the causal relationships for the whole gene set analysis, as it is too time-consuming. We believe the performance will be similar to the selective analysis.

#### Results of OpenTargets enrichment analysis

In this study, we tested whether our proposed method could identify potential targets (as listed in the OpenTargets database) based on the overall association scores. In the OpenTargets database, the overall association score is an aggregated target-disease association evidence that combines 7 different association evidence, including genetic association, somatic mutation, known drug, affected pathway, RNA expression, literature support, and animal model. We tried 10 different association score cutoffs between 0 and 1 with a step size of 0.1 to define real targets. Meta-analysis was performed using harmonic mean p-value (HMP) to combine p-values derived under different score cutoffs. Briefly, HMP^34^ is a meta-analysis method based on Bayesian model averaging, which combines p-values derived from dependent hypotheses while appropriately controlling for family-wise error rate (FWER). Table 9 summarizes the enrichment analysis results. As can be seen from the table, the identified directly causal genes for 6 binary traits were found to be significantly or nominally enriched for their corresponding targets. For the remaining 5 binary traits (anxiety, COVID-19, depression, heart failure, stroke), the number of identified directly causal genes in relevant tissues was no more than 3. Unsurprisingly, it’s difficult to find potential targets from such a small set of candidates, which unavoidably limit the power of the fisher’s exact test in detecting significant differences. This problem could be alleviated when the available samples increase. Overall speaking, our proposed method could identify potential targets for studied disorders. These enrichment analysis results further verified the validity of our proposed framework in identifying causally relevant genes for different traits. For more details about the enrichment analysis results under different cutoffs, please refer to Table S9.

In addition, we examined whether our proposed method could identify more potential targets than the conventional univariate-based analysis. Since the sample size has a tremendous effect on the power of the fisher’s exact test in detecting differences, in this study, we only consider tissue-specific traits with >= 5 directly causal genes. Tables 10 and S10 summarize the comparison results. For all studied binary traits with >= 5 causal genes, our method was able to identify significantly more targets than the univariate test. Most importantly, it consistently achieved superior performance than the commonly used univariate test in identifying potential targets regardless of the number of directly causal genes (Table S10.b), which is of great clinical significance. Furthermore, we tested whether our proposed method could identify more targets than the univariate test under the same p-value cutoff, i.e., p=0.001. Even though the univariate test identified much more trait-associated genes, our method still achieved better performance in identifying potential targets for most disorders (Table S10.c and S10.d). In this regard, these identified core genes, especially these novel genes (Table S11), have the potential to serve as targets for new drugs and therapeutic treatments for studied disorders.

Table 11 demonstrates the identified novel genes that were top ranked (z score) in our inferred causal graphs for the studied binary traits. As shown in the table, our method identified PIGIS as a novel core gene for CAD in the artery coronary. In a previous study, Yi et al. found it played a significant role in the regulation of blood flow and platelet activity, which could lead to the development of atherothrombosis and acute coronary syndromes^35,36^. FXYD2, a top core gene identified from our method for diabetes in the pancreas tissue, was proven to be intensively involved in the Akt pathway^37^, which controls pancreatic beta cell growth and proliferation. For breast cancer in the breast mammary tissue, ATXN3 was one of the top ranked core genes identified by our method. Through deubiquitinating KLF4, it could accelerate the metastasis of breast cancer ^38^. The comparison results further corroborated the validity and superiority of our proposed method in identifying disease-related genes.

#### Results of survival and replication analysis for breast cancer

We separately performed survival analysis on gene sets identified from the breast mammary and whole blood tissues. For the breast mammary tissue, we identified 7 directly causal genes for breast cancer. As can be seen from Fig.6, 6 of them were found to be influential on the survival of breast cancer patients. For the whole blood tissue, we identified 8 genes. All 8 genes have a profound effect on the survival of patients. Table S12 demonstrates the survival analysis results for gene sets identified from our method and that from the univariate-based analysis. Almost all the genes identified from both methods play essential roles in the survival of breast cancer patients. Overall, our method achieved better performance with a much lower combined p-value (3.9E-12) than the univariate test (2.33E-11) based on the Simes method. These results provide further support for the validity and efficacy of our proposed framework in identifying disease-related genes.

**Fig. 6.**
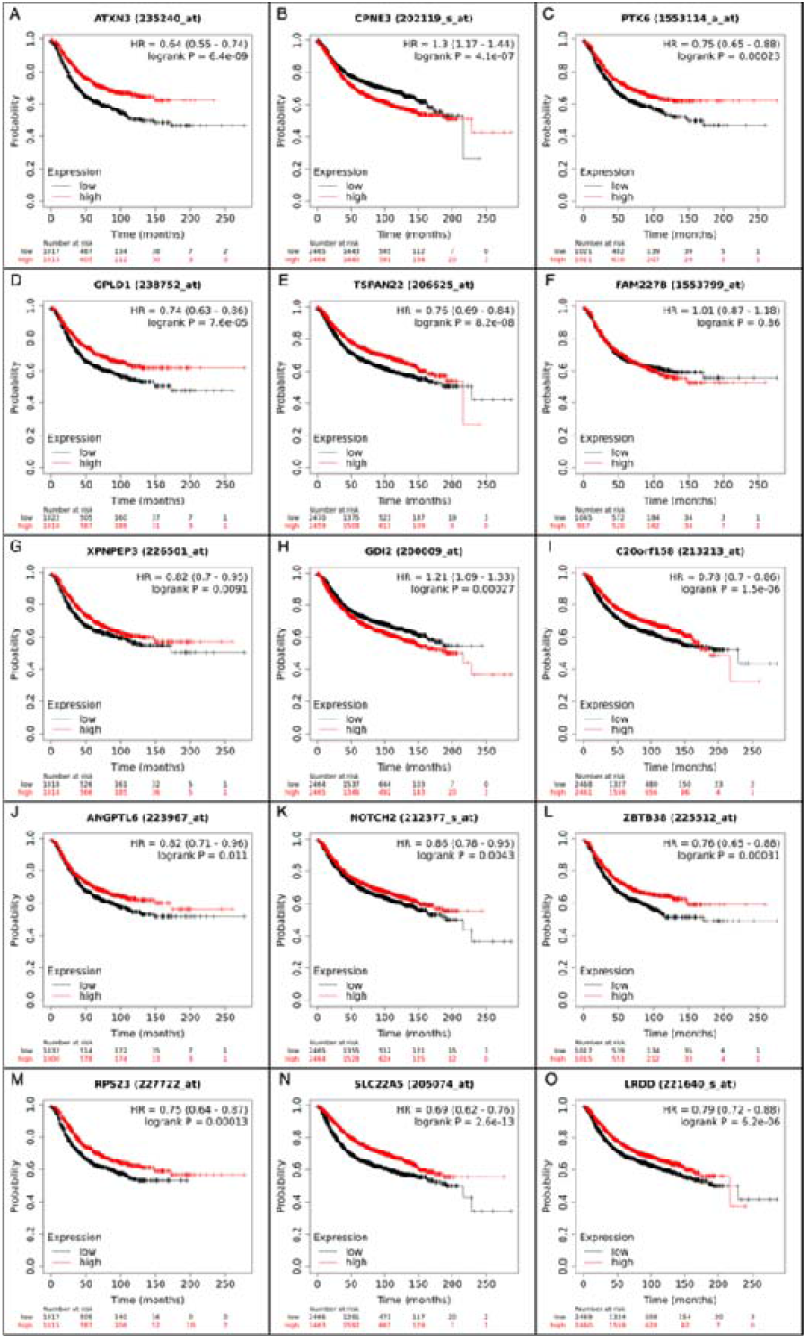
Survival analysis of directly causal genes identified from the breast mammary(A-G) and whole blood(H-O) for breast cancer.

Some of the identified core genes, for example NOTCH2, PTK6, were also known pathogenic/likely pathogenic genes for breast cancer based on the OncoVar database. Most importantly, the proportion of identified pathogenic genes were significantly higher than expected with a p-value of 0.037. According to the OncoVar, 3 of the identified core genes, i.e., NOTCH2, SLC22A5, PTK6, were drug-related genes for breast cancer. All of them were known mutation associated genes. The replication analysis further demonstrates the reliability of our proposed method in identifying core genes for studied trait.

#### Results of multiple traits analysis

Two- and three-traits analyses were performed to demonstrate the feasibility of our proposed framework in identifying causally relevant genes for multiple traits. To be more specific, we performed 6 two-traits and 2 three-traits analyses. Table 12 demonstrated exposure and outcome traits for multiple traits analyses as well as the identified causally relevant genes. In this study, we inferred the causal graphs for multiple traits involving BMI, HbA1c, LDL as exposure traits, and CAD, diabetes, COVID-19 as outcome traits. Many studies have revealed the causal relationships among our studied traits^39-44^. As expected, the inferred causal graphs for multiple traits were much more complex than those for single traits. All inferred causal graphs had a high proportion of indirect causal genes, with some even reaching 90%. Take CAD for example, when we performed a single trait analysis in the whole blood tissue, the proportion of indirect causal genes was 30.7%. If we incorporated other risk factors like BMI and LDL, the proportion of indirect causal genes increased to around 90%. Given this, we could conclude that while some genes directly affect the development of a particular disease, a large proportion of genes probably contribute to the development of disease through their effects on different risk factors. We also compared the direct causal genes identified from single trait with that from multiple traits analyses for the same outcome trait (Fig.7 and Table 13). Even though the proportion of direct causal genes dramatically changed, the identified causally relevant genes were comparatively stable. In other words, many identified core genes remained stable regardless of the existence of other risk factors. Contrary to this, some identified core genes in the single-trait analysis became peripheral after incorporating other risk factor traits into the analysis. Take UHRF1BP1 for example, it was identified as a core gene for CAD in the whole blood tissue. However, after we incorporated BMI into the analysis, it became an indirect causal gene. We hypothesized that UHRF1BP1 might exert its effect on CAD through obesity. Nikpay et al. ^45^ verified this hypothesis in their recent work. Compared with single-trait analysis, core genes identified from multiple traits analysis were inclined to have multiple action paths. As shown in Table 12, 5 of the 14 core genes identified for CAD (incorporating BMI and diabetes as risk factors) could exert their effects through other risk factors. However, for single-trait analysis, none of the identified core genes were found to have additional action paths to CAD.

**Fig. 7.**
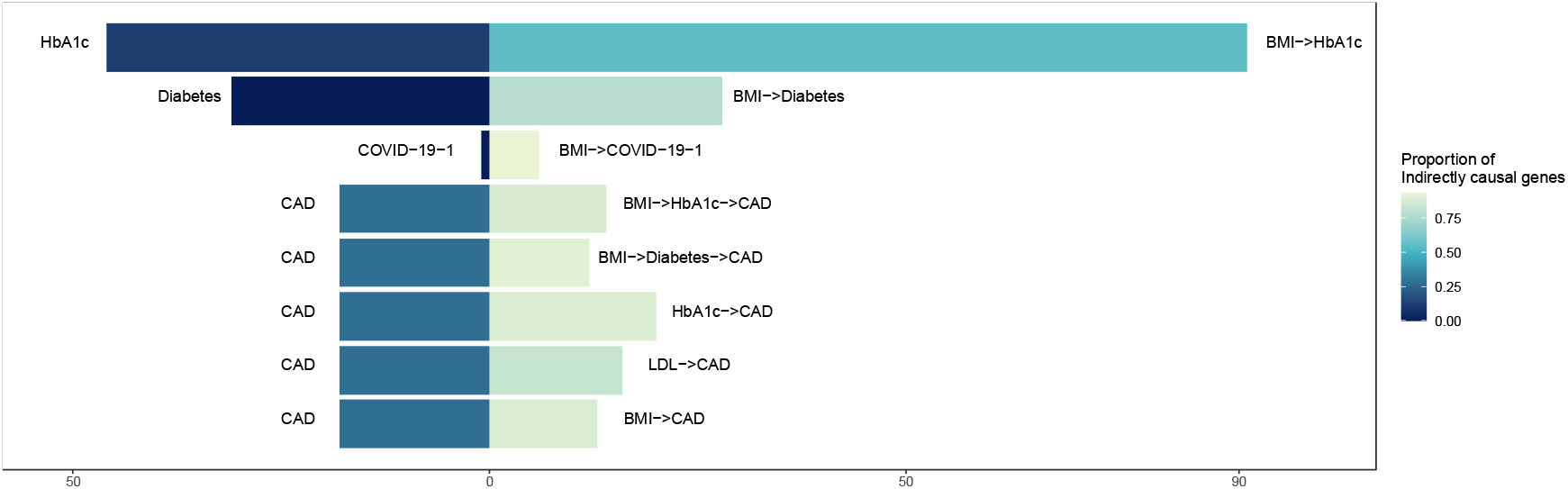
Comparison of identified directly causal genes and proportion of indirectly causal genes between multiple and single trait analysis.

We also estimated the total and direct causal effects of different genes on the “outcome” traits for all multiple traits analysis scenarios (Table S13). As can be seen from Table 14 and S13, most identified core genes had strong effect sizes. Notably, these genes were also identified as significant core genes in the single-trait analysis. Most importantly, the estimated causal effects of these genes on the studied disease were very stable. They barely changed even we included other risk factors into the analysis. The effect size of some core genes may change with the risk factors. Even so, the directions of the estimated causal effects were usually consistent. Notably, many peripheral genes also had significant influences on the studied disorders. Compared to single-trait analysis, the proportion of peripheral genes for multiple traits analysis has substantially increased. The estimated total and direct causal effects could help us better understand the contribution of corresponding genes to the development of the disorders, which may shed light on the discovery of new targets for therapeutic treatment.

#### Results of additional analyses

Table S14 summarizes the enriched pathways for all studied traits. As expected, most of the enriched pathways were associated with the studied traits or related pathophysiology. Different researchers have studied their roles in the corresponding traits. Some of them were even identified as new therapeutic targets. For example, NR1H3 & NR1H2(LXRs) regulate gene expression linked to cholesterol transport and efflux was the top enriched pathway for CAD in the artery coronary tissue. These LXRs (Liver X receptors) were suggested to be intensively involved in the efflux, transport, excretion of cholesterol and lipid in different cell types and responsible for the development of CAD ^46-48^. Also, they had direct effects on the development of cardiovascular disease. In a previous study, Cannon et al. suggested that activation of LXRs could lead to cardiomyocytes damage and loss ^49^. Other researchers have proven that synthetic LXRs agonists are effective in inhibiting the development of atherosclerosis^46^. Take LDL in the whole blood tissue as another example, Lysosome Vesicle Biogenesis, and Lysosome were the top 2 enriched pathways. They were both intimately connected to the biogenesis and transport of lipid^49^. For more details about the enriched pathways, please refer to Table S14.

Table S15 demonstrates the drug enrichment analysis results of identified core genes for our studied binary traits. Some of the enriched drugs have been tested/used for the treatment of patients in clinical practice. For example, dasatinib was significantly enriched for breast cancer in the breast mammary tissue. In a related study, Tian et al. demonstrated that dasatinib combined with paclitaxel could inhibit tumor growth in triple-negative breast cancer patients^50^. For COVID-19 in the lung, Niclosamide was the top enriched drug. There is abundant evidence suggesting the potential clinical usage of this drug ^51-53^. Also, Tufts Medical center has an ongoing clinical trial to evaluate whether it could be repurposed for the treatment of mild/moderate COVID-19 patients ^54^.

Also, we analyzed the enriched transcription factors for different traits. Again, most of the enriched transcription factors were intimately connected to the underlying pathophysiology of the corresponding disorders. For example, BHLHA15, which was significantly enriched for the diabetes, was proven to be essential in maintaining the secretory cell architecture ^55^. For more details on the enriched transcription factors, please refer to Table S16. These enrichment analysis results provide further support for the validity and efficacy of our proposed method in identifying core genes for different traits.

In this study, we compared the “core” genes and “peripheral” genes as well as their estimated effects from our proposed method to the susceptible genes identified from the univariate test (Table S5). Our proposed method tended to identify fewer genes than the univariate test when the p-value cutoffs were all set to 0.001. This was expected due to the more stringent criteria of our proposed method. For some trait-associated genes, the estimated effects from both methods were very close. For example, the estimated effects of LPL on CAD in the whole blood tissue from both methods were approximately equal. Previously, Khera et al. ^56^ reported its significant role in the clearance of triglycerides, which could lead to the onset of CAD. Based on the comparison result, we assumed that its function in the regulation of triglycerides levels probably was not influenced by other CAD-associated genes. However, there also existed many genes for which their estimated effects from the two methods were quite different. Take SLC22A5 for example, the estimated effect on breast cancer in the whole blood tissue from the univariate test was obviously stronger than that from our method. It has been proven to have a critical role in regulating breast cancer cell proliferation. Wang et al. demonstrated that the expression of SLC22A5 in breast cancer patients was induced by estrogen signaling, which was regarded as the most critical factor for breast cancer ^57^. In other words, other causally relevant genes could regulate its expression. The IDA and jointIDA methods take the influence network into account while estimating the effects of the identified gene. Thus, the estimated effects were less likely to be biased by other factors. Even though the estimated effects from the two methods were not always consistent, the directions of the effects were generally concordant. For more detail about the comparison results, please refer to Table S5.

External knowledge could also be incorporated while estimating causal graphs for different traits. In this study, we explored the usage of external knowledge on gene-gene interaction in the whole blood tissue for causal graph estimation. As we only included external knowledge between different genes, the inferred gene-outcome causal networks shall remain unchanged. Genes associated with the studied trait were of primary interest and clinical significance, thus we only analyzed those trait-associated genes. Table S17 summarized the estimated causal graphs for various traits in the whole blood tissue after incorporating external knowledge. As expected, the incorporated external knowledge only mildly affected our estimated causal graphs.

Univariate-based analyses were also performed for all the multiple traits analysis scenarios. All the genetically predicted traits were regarded as “genes” while performing the univariate test. Table S13 summarizes the comparison results of the estimated causal effects from our method and that of the univariate test. Again, our method identified fewer trait-associated genes (here we mainly refer to core genes) than the univariate test. While the directions of the estimated effects from both methods were consistent with each other, the difference between estimated effects varied for different genes. For many identified core genes, the estimated total causal effects were very close to the effects derived from the univariate test. However, there still existed some cores for which the estimated effects from our method were obviously different from that of the univariate test.

Our proposed method was able to differentiate core genes from peripheral ones. Most importantly, it could eliminate spurious associated genes confounded by actual core genes. However, if the univariate test was utilized, then both the indirectly causal genes and non-causal genes confounded by real core ones will be identified as susceptible genes for our studied traits. As can be seen from Tables S5 and S13, for most studied traits, more than half of the identified susceptible genes were not identified as causal based on our method. While the univariate test was able to identify more susceptible genes, most of them were in fact not related to the studied traits. Thus, they were unlikely to be the potential targets for the therapeutic treatment of the corresponding disorder. Apart from the ability to qualify the causal relationships between genes and studied traits, our method could also quantify the total and direct effects of identified genes on different traits. Compared with the univariate test, our method was less biased in estimating the effects of trait-associated genes due to its ability to eliminate confounding effects. Since many genes may have more than one action path to our studied traits, quantifying both the direct and indirect causal effects could help us comprehensively understand their influence on the development of certain diseases. Most importantly, these identified causal genes as well as the estimated causal effects may shed light on prioritizing new targets for therapeutic treatment.

## Discussion

This study proposed a novel framework capable of identifying different types of causal genes and quantifying their total and direct causal effects on different traits. Simulation results demonstrated the validity and efficacy of our proposed method in uncovering causal relevant genes and estimating their causal effects. It is worth noting that it consistently achieved superior performance than the commonly used gene-based analysis (univariate test). Besides, we validated the efficiency and robustness of our proposed method in identifying causally relevant genes by split-half replication analysis. The replication rates for most continuous traits were rather good. Due to the complex nature of most complex diseases (binary traits), the replication rates for them were less compelling. Despite this, we can still replicate genes with large effects on the studied disorders. Stability selection further reflects the reliability of our proposed method in inferring the core genes as well as the overall causal structure of studied traits. It achieved satisfactory performance in qualifying the causal genes and quantifying their causal effects with no inflated type I errors. Most importantly, our method could identify more drug targets than the commonly used gene-based analysis, which is also based on imputed expression profiles. Usually, the univariate test identified more trait-associated genes than our proposed method. However, our proposed method achieved better performance in identifying potential targets for most disorders. Survival analysis further revealed the superiority of our proposed method in identifying survival-related genes for breast cancer.

The presented framework has several strengths. A distinct advantage is that it’s a unified framework that can estimate the overall causal structure of studied traits from the GTEx and GWAS-imputed gene expression profiles. In other words, it allows the overall causal structure to be estimated from different samples, which dramatically improves the flexibility of the proposed method. While the mediator genes affect the studied traits through other genes, the core genes directly contribute to the development of the corresponding disease or trait variation. Thus, we could gain a better and more comprehensive understanding of the underlying biological mechanisms for different traits. Most importantly, the identified cores genes for these disorders have the potential to serve as targets for new drugs and therapeutic treatments. In addition, we have proposed a novel p-value based regularization method for causal effects estimation. It enables us to quantify the total, direct and mediating causal effects of identified causally relevant genes on studied traits based on imputed expression profiles with satisfactory accuracy. The estimated causal effects could reflect the “importance” of these identified genes to studied traits, which could shed light on the prioritization of targets for treatment. Since the expression profiles were imputed from genotypes, the identified causally relevant genes as well as their causal effects are unlikely to be confounded by other factors such as medication usage. Furthermore, it can be potentially extended to more than one trait. While we focused on expression data in this study, the framework is general and can be applied to methylomic, proteomic, and other kinds of omics data to decipher their causal effects. Apart from this, external knowledge could be incorporated into the estimated causal networks.

Several methods have been developed to identify genes linked to complex traits based on GWAS. However, their focus and methodology were different from the current work. For example, SMR was developed to combine GWAS and eQTL data to prioritize genes linked to a trait/disease based on principles of MR. Some top associated cis-eQTL could be in LD with two distinct causal variants, one affecting gene expression and the other affecting the trait of interest. To distinguish from pleiotropy (the same cis-eQTL affecting both expression and trait), Zhu et al. ^58^ also developed the HEIDI test. Another closely related line of work is to ‘impute’ expression based on a reference dataset (e.g., GTEx), also known as TWAS (transcriptome-wide association study). Please refer to ^59^ for details.

The current work is different from the above in several ways. Firstly, one of our main aims is to produce a directed network uncovering links between genes and phenotypes, whereas the above methods focus on univariate associations. Here we present a unified framework that enables the total, direct and mediating effects of a single or any set of genes to be estimated, building on the principles of the Bayesian network and do-calculus for causal inference. We also propose ways to extend the approach to incorporate external knowledge (e.g., other pathways or protein-protein interaction databases), and analyze more than one clinical trait/disease.

The methods described above are largely based on the principles of MR, where the eQTL(s) (or other types of QTLs) serve as instruments. The presented framework also involves estimating expression from eQTLs, which share the advantages of MR, but we also model other genes in a BN and estimate causal effects by accounting for other genes. A distinct advantage is that we may also model the effect of mediating genes and direct causal effects, hence gaining insight into the mechanisms underlying the gene’s effect on the phenotype(s). For example, gene A may exert its effect on the outcome via gene B, i.e., gene B is a mediator. The total causal effect of A and causal mediation effect via B can be obtained under the current framework, but not by existing methods like SMR or TWAS.

In addition, in MR (on which most current methods are based) a key assumption is that the instruments should be free from (imbalanced) horizontal pleiotropy, i.e., the instrument SNP(s) should not have its effects mediated through other pathways other than the exposure. It is hard to test this assumption in expression-based MR studies as the number of instrumental SNPs is often small. Also, SNPs in LD with the instrument SNP(s) can also have pleiotropic effects. While the HEIDI test can partially ameliorate the latter issue (also referred to as ‘linkage’), it has its own limitations. For example, HEIDI was not designed for multiple causal variants at the locus, and pleiotropy and linkage cannot be distinguished. Here we tested for the association of each gene with the outcome, controlling for all possible subsets of other genes. A SNP can affect the outcome via other pathways other than through the exposure, but it is reasonable to believe that biologically it is common (although not always) that the instrumental SNP exerts its pleiotropic effect on the outcome via expression changes of some other genes (+/- other downstream pathways or risk factors). By controlling for other genes in our model, we also reduce the chance of pleiotropic pathways affecting our result. The principle holds no matter whether the instrumental SNPs themselves exert pleiotropic effects or they are in LD with other variant(s) which show horizontal pleiotropy.

Another assumption of MR is that the instruments are not associated with measured or unmeasured confounders. In ordinary MR, it is possible that the SNP instruments, or those in LD with them, are associated with confounders such as other genes (e.g., a gene may lead to both expression changes in the studied gene and the outcome). Again, by controlling for expression of other genes, the risk of confounding can be reduced. Our approach is similar to multivariable MR (MVMR), but here we controlled for all possible subsets of other ‘exposures’(genes) instead of performing a single MVMR. MVMR has been used to control for confounders that may violate MR assumptions ^60^. The above study also shows that MVMR gives correct estimates of the direct causal effect no matter whether the covariates are confounders, mediators, or colliders.

Bayesian network (BN) has been used to infer gene regulatory networks and has been employed in several works ^61^. Nevertheless, the main applications of BN were mainly concentrated on microarray/RNA-seq data. A limitation is that typically only small sample sizes are available for a given disease, especially if the tissue of interest is hard to access (e.g., the brain). In addition, reverse causality is a concern as expression changes may be a result of the disease or medications. As a result, building a network integrating both gene-gene connections and clinical phenotypes is not easy. The presented framework allows gene expression and disease trait(s) to be estimated in different samples, greatly improving the flexibility and applicability of the approach.

There are a few limitations in this study. The correction estimation of IDA/jointIDA requires several assumptions, including that the variables should follow a multivariate normal distribution, and that the distribution of the variables should be faithful to the true underlying DAG. The latter (faithfulness assumption) is satisfied if the data is generated from a DAG with no hidden confounding variables. In practice, it is almost impossible to guarantee the lack of hidden confounders. Here we model the effects of genes on diseases/traits similar to a two-stage least squares (2SLS) approach in instrumental regression, in which predicted expressions were used, which reduces the risk of confounding. We note that the instrumental SNPs may be associated with (unknown) confounders, which is a limitation. However, we did not model one gene at a time but also considered a number of other genes. As explained earlier, the inclusion of other genes further reduced the risk of confounding as their effects can be controlled for. Another assumption is that the graph is acyclic, which is likely violated in real biological networks where feedback loops are often present.

We performed two sets of analyses. The first is restricted to around a hundred and fifty (or less) genes that are selected by PC-simple. In other words, we restricted the analysis to a set of genes with directly causal effects. With a smaller number of genes to model, the network can be constructed more accurately and visualized more readily. Moreover, it took less time to decipher the network and quantify the causal effect of each gene. A drawback is that there may be hidden confounders not modeled. Also, the genes selected by PC-simple are likely to have a relatively strong direct effect on the outcome, while genes with indirect causal effects (mediated by other genes) are unlikely to be included and modeled. We also conducted an analysis that includes a much large number of genes (∼ 3000) whose expression can be imputed. Based on ^62^, it was observed that IDA could discover top causal genes from expression studies much better than by chance and other methods with a large number of features (5,361) and a very modest sample size (N=63). The number of genes is smaller in our case, but the sample size is around 10 times higher. Although we could not verify our approach directly by simulations when the number of genes is large (e.g., >1000) due to computational burden, we believe the method can perform reasonably well in identifying the top causal genes. The entire gene network may not be estimated very accurately compared to the case when the gene number is small, but IDA or jointIDA methods only require the local neighborhood of the network to be estimated accurately. When we model a larger number of genes, an advantage is that the risk of hidden confounders is further reduced.

Genes whose expression cannot be reliably imputed are not included in estimating causal effects, but we argue that they are unlikely to be major confounders. This is because our assumption requires the instrumental SNPs to be uncorrelated with confounders, but genes without reliable imputed expressions are unlikely to have strong associations with existing eQTLs (i.e., the instruments).

As shown in our simulations, a larger sample leads to a more accurate estimation of causal effects. The current sample size of GTEx is still relatively modest, but larger eQTL consortiums (e.g., https://www.eqtlgen.org/) are becoming available ^63^.

With the above limitations, we consider our framework to be a way to prioritize genes with high causal or direct causal effects, and as a way to estimate the ‘variable importance’ of such genes.

## Data Availability

The UK Biobank data is available to qualified researchers upon application.

## Data availability

Relevant R code will be available on github on or before publication.

## Acknowledgements

This work was supported partially by a National Natural Science Foundation China (NSFC) Young Scientist Grant (31900495), a National Natural Science Foundation China grant (81971706), the Lo Kwee Seong Biomedical Research Fund from The Chinese University of Hong Kong and the KIZ-CUHK Joint Laboratory of Bioresources and Molecular Research of Common Diseases, Kunming Institute of Zoology and The Chinese University of Hong Kong, China.

