## Supplementary material for "A Bayesian network-based framework to uncover the causal effects of genes on complex traits based on GWAS data": Supp Text

**Supplementary text**

**Gene-phenotype causal network inference**

The imputation for gene expression levels is best conducted on a full set of [genetic variants](https://www.sciencedirect.com/topics/biochemistry-genetics-and-molecular-biology/genetic-divergence) instead of SNPs on the genotyping panel only. Therefore, we firstly employed the program Minimac using the University of Michigan Imputation Server and 1000 Genomes Phase 3 v5 as the reference panel to implement the variant-level imputation. SNPs with INFO score >0.3 were kept. We then employed PrediXcan to impute expression levels from the imputed genotype data. Briefly, the algorithm first produces prediction models for expression levels from an external reference dataset (such as GTEx) which contains both genotype and expression data. An elastic net regression model is used by default. Then, the prediction model can be applied to independent genotype data to impute gene expression data. Expression in different tissues can be estimated as long as the reference dataset contains such data.

**PC-simple algorithm**

Briefly, PC-Simple can be regarded as a generalization of correlation screening that utilizes ordered independence screening to infer the causal relationships between covariates and response.

To start with, we need to define a few notations. For a set $S\subseteq\{1,2,\ldots,p\}$, $\left| S \right|$ is the cardinality of the set, $S^{C}$ defines its complement in $\{1,2,\ldots,p\}$. Besides, $\rho(Z^{1},Z^{2}|W)$ and $parcov(Z^{1},Z^{2}|W)$ respectively indicate the partial correlation and covariance between two variables given variable(s) $W$. Let $X=[X^{1},X^{2},\ldots X^{p}]$ be a $n\times p$ matrix of adjusted gene expression data for *p* genes, *Y* be a vector of the corresponding adjusted phenotype dataset for *n* subjects. Suppose *Y* is defined by the following linear regression model:

$$Y= \sum_{j=1}^{p} \beta^{j}X^{j}+\varepsilon$$

(1)

Here, $\varepsilon$ indicates the noise item that is independent of $X^{j}$ and follows a multivariate normal distribution ($\epsilon\sim N(0,\sum)$). For this gene-phenotype generative model, we consider most or some of the $\beta^{j}$ are zero, while the remaining are nonzero for the studied phenotype. Our goal here is to identify the active gene set $G=\{j=1,2,\ldots,p;\beta^{j}\neq0\}$. Under the partial faithfulness assumption, we have that:

$$\rho\left( {Y,X}^{j} | X^{S} \right)\neq0 for all S\subseteq\left\{ j \right\}^{C} if and only if\beta^{j}\neq0$$

(2)

In other words,

$$\rho\left( {Y,X}^{j} | X^{S} \right)=0 for some S\subseteq\left\{ j \right\}^{C} implies\beta^{j}=0$$

(3)

To identify the active gene set, we firstly set the conditional set $S= \emptyset$. Then we could get the first candidate active gene set by screening all marginal correlations between pairs $\left( Y,X^{j} \right), for j=1, 2,\ldots p$, i.e.,

$$G^{1}=\{j=1,2,\ldots,p;cor(Y,X^{j})\neq0\}$$

Following this step, we could identify the second candidate active gene set through performing partial correlation screening based on expression (2).

$$G^{2}=\{j \epsilon G^{1};\rho(Y,X^{j}|X^{k})\neq0 for all k \epsilon G^{1}\backslash\{j\} \}\subseteq G^{1}$$

Through recursively performing partial correlation screening with increased order of conditional set based on expression (2), we could exclude genes from previous candidate active gene set until it does not change anymore. In this study, the recursive formula was employed to estimate the partial correlation:

$$\hat{\rho}\left( Y,X^{j} | X^{S} \right)= \frac{\hat{\rho}(Y,X^{j}|X^{S}\backslash\{X^{k}\})-\hat{\rho}(Y,X^{k}|X^{S}\backslash\{X^{k}\})\hat{\rho}(X^{j},X^{k}|X^{S}\backslash\{X^{k}\})}{{[\{1-{\hat{\rho}(Y,X^{k}|X^{S}\backslash\{X^{k}\})}^{2}\}\{1-{\hat{\rho}(X^{j},X^{k}|X^{S}\backslash\{X^{k}\})}^{2}\}]}^{1/2}}$$

(4)

Specifically, the nth order partial correlation could be computed from three (n-1)th-order partial correlations. To improve the computational efficiency of this algorithm, we set the maximum order for partial correlation screening to be 3. In other words, those genes that survived the 3-order partial correlation screening were deemed to be causal for the studied phenotype. Fisher’s Z-transform was employed to test whether a partial correlation is zero, i.e.,

$$Z\left( Y,X^{j} | X^{S} \right)= \frac{1}{2}\{\frac{1+\hat{\rho}\left( Y,X^{j} | X^{S} \right)}{1-\hat{\rho}\left( Y,X^{j} | X^{S} \right)}\}$$

(5)

As suggested by Buhlmann et al ^10^, the null hypothesis $\hat{\rho}\left( Y,X^{j} | X^{S} \right)=0$ would be rejected if ${(n-\left| S \right|-3)}^{1/2}\left| Z\left( Y,X^{j} | X^{S} \right) \right|>\phi^{-1}(1-\alpha/2)$. Here $\alpha$ and $\phi$ respectively denote the significance level and standard normal cumulative distribution function. In this study, we set $\alpha=0.001$. After employing the PC-Simple algorithm, we could get the causal relationship between genes and clinical phenotypes, with a vector $Z_{p\times1}$ indicating the reliability of the inferred causal relationships. Notably, we could convert the Z score vector into a p-value vector by: $P=2*\phi(-abs(Z))$.

IDA method

For a given causal graph $G$, the distribution generating the graph can be represented as follows:

$$f\left( X^{1},X^{2},\ldots X^{p},Y \right)=f(Y|{pa}_{Y})\prod_{i=1}^{p} f(X^{i}|{pa}_{i})$$

(6)

Here ${pa}_{i}$ indicates the parents nodes set for variable $X^{j}$, similarly for ${pa}_{Y}$. The distribution for target $Y$ generated by an intervention on $X^{i}$ could by summarized as:

$$E\left( Y | do\left( X^{i}=x_{i}^{'} \right) \right)=\left\{ \begin{aligned} E\left( Y \right), if Y\in\text{Pa}\text{i} \\ \int E\left( Y | x_{i}^{'},{pa}_{i} \right)f\left( {pa}_{i} \right)d{pa}_{i}, if Y\notin{pa}_{i} \end{aligned} \right.$$

(7)

The causal effects of node $\boldsymbol{X}^{\boldsymbol{i}}$ on target $Y$ can be estimated by intervention calculus as follows:

$$\frac{\partial}{\partial x}E\left( Y | do\left( X^{i}=x_{i}^{'} \right) \right)=E\left( Y | do\left( X^{i}=x^{i}+1 \right) \right)-E\left( Y | do\left( X^{i}=x^{i} \right) \right)$$

(8)

Under linear assumption, the total causal effects of $X^{i}$ on Y is the regression coefficients of $X^{i}$ in the regression of Y on $X^{i}$ and${pa}_{i}$ with $Y\notin{pa}_{i}\text{i}$

jointIDA method

jointIDA is an extension of this method with multiple simultaneous interventions. Similar with IDA, the total joint effect of ${(X}^{1},X^{2},\ldots X^{k})$on $Y$ can be given by:

$$\theta_{iY}^{(1,2,\ldots k)}:=\frac{\partial}{\partial x^{i}}E\left( Y | do\left( {(x}^{1},x^{2},\ldots x^{k} \right) \right), for i=1,2,\ldots k$$

(9)

Under linear assumption, the total joint effect can be interpreted as follows:

| $\theta_{iY}^{(1,2,\ldots k)}=E[\left( Y \vert do\left( {(x}^{1},x^{2},x^{i}+1,\ldots x^{k} \right) \right)]$ *-* $E\left[ \left( Y \vert do\left( {(x}^{1},x^{2},x^{i},\ldots x^{k} \right) \right) \right]$ | (10) |
| --- | --- |

When the causal graph is known, we could identify paths from node $\boldsymbol{X}^{\boldsymbol{i}}$ to the target $Y$. Based on the recursive regressions, the total joint effect can be given as follows:

$$\theta_{iY}^{[k]}=\theta_{iY}^{[k]\backslash\{j\}}-\theta_{ij}^{[k]\backslash\{j\}}\theta_{jY}^{[k]\backslash\{i\}} for any j\in\{1,2,\ldots,k\}\backslash\{i\}$$

(11)

Here $\left[ k \right]\mathrm{and}[k]\backslash\{j\}$ denote $(1,2,\ldots,k)$ and $(1,\ldots,j-1,j+1,\ldots k)$ respectively. If we intervene on $\boldsymbol{X}^{\boldsymbol{i}}$ and all parents nodes set of the target $Y$ ($Pa(Y))$, then the estimated causal effect for intervention node $\boldsymbol{X}^{\boldsymbol{i}}$ is the direct causal effect.
