## Supplementary Tables/Figures for "A Bayesian network-based framework to uncover the causal effects of genes on complex traits based on GWAS data"

Table S1 Parameters settings for simulation scenarios

| No. of Genotypes | No. of Genes | No. of Overall samples | No. of GTEx Samples | Graph density | Min weight in graph |
| --- | --- | --- | --- | --- | --- |
| 500 | 50 | 100000 | 800 | 0.05 | 0.1 |
| 500 | 50 | 100000 | 800 | 0.05 | 0.3 |
| 500 | 50 | 100000 | 800 | 0.05 | 0.5 |
| 700 | 70 | 100000 | 800 | 0.05 | 0.1 |
| 700 | 70 | 100000 | 2000 | 0.05 | 0.1 |
| 700 | 70 | 100000 | 8000 | 0.05 | 0.1 |
| 700 | 70 | 100000 | 800 | 0.05 | 0.3 |
| 700 | 70 | 100000 | 2000 | 0.05 | 0.3 |
| 700 | 70 | 100000 | 8000 | 0.05 | 0.3 |
| 700 | 70 | 100000 | 800 | 0.05 | 0.5 |
| 700 | 70 | 100000 | 800 | 0.1 | 0.1 |
| 700 | 70 | 100000 | 800 | 0.1 | 0.3 |
| 700 | 70 | 100000 | 800 | 0.1 | 0.5 |
| 700 | 70 | 100000 | 800 | 0.02 | 0.1 |
| 700 | 70 | 100000 | 800 | 0.02 | 0.3 |
| 700 | 70 | 100000 | 800 | 0.02 | 0.5 |
| 1000 | 100 | 100000 | 800 | 0.05 | 0.1 |
| 1000 | 100 | 100000 | 800 | 0.05 | 0.3 |
| 1000 | 100 | 100000 | 800 | 0.05 | 0.5 |
| 1500 | 150 | 100000 | 800 | 0.05 | 0.1 |
| 1500 | 150 | 100000 | 800 | 0.05 | 0.3 |
| 1500 | 150 | 100000 | 800 | 0.05 | 0.5 |

*Maximum Weight in graph = Min Weight*1.2

Table S2 Outlier analysis for correlation of 20 replicates

| Replicates_Index | Correlation | Initial_Est | Adj-pvalue | Adj-pvalue-i | Univariate test | Outlier |
| --- | --- | --- | --- | --- | --- | --- |
| 1 | CorIDA | 0.420 | 0.750 | 0.823 | 0.420 |  |
| 2 | CorIDA | 0.527 | 0.554 | 0.593 | 0.527 |  |
| 3 | CorIDA | 0.709 | 0.917 | 0.919 | 0.709 |  |
| 4 | CorIDA | 0.006 | 0.783 | 0.791 | 0.006 |  |
| 5 | CorIDA | 0.249 | 0.244 | 0.244 | 0.249 | * |
| 6 | CorIDA | 0.754 | 0.817 | 0.832 | 0.754 |  |
| 7 | CorIDA | 0.839 | 0.878 | 0.887 | 0.838 |  |
| 8 | CorIDA | 0.706 | 0.729 | 0.740 | 0.705 |  |
| 9 | CorIDA | 0.577 | 0.857 | 0.927 | 0.578 |  |
| 10 | CorIDA | 0.175 | 0.207 | 0.433 | 0.176 | * |
| 11 | CorIDA | 0.636 | 0.673 | 0.726 | 0.636 |  |
| 12 | CorIDA | 0.617 | 0.891 | 0.914 | 0.616 |  |
| 13 | CorIDA | 0.755 | 0.792 | 0.897 | 0.755 |  |
| 14 | CorIDA | 0.819 | 0.876 | 0.907 | 0.819 |  |
| 15 | CorIDA | 0.737 | 0.820 | 0.826 | 0.737 |  |
| 16 | CorIDA | 0.725 | 0.833 | 0.852 | 0.725 |  |
| 17 | CorIDA | 0.766 | 0.815 | 0.866 | 0.767 |  |
| 18 | CorIDA | 0.788 | 0.801 | 0.808 | 0.786 |  |
| 19 | CorIDA | 0.289 | 0.571 | 0.589 | 0.289 |  |
| 20 | CorIDA | 0.782 | 0.799 | 0.802 | 0.782 |  |
| *Parameter Setting: SNPs700/Genes70/OverallSamples100000/GtexSamples800/GraphDensity0.05/Weight0.1 | | | | | | |

Table S3 Outlier analysis for RMSE of 20 replicates

| Replicates_Index | RMSE | Initial_Est | Adj-pvalue | Adj-pvalue-i | Univariate test | Outlier |
| --- | --- | --- | --- | --- | --- | --- |
| 1 | RMSE_IDA | 2.252 | 0.967 | 0.761 | 2.251 |  |
| 2 | RMSE_IDA | 1.090 | 0.957 | 0.830 | 1.088 |  |
| 3 | RMSE_IDA | 1.261 | 0.656 | 0.650 | 1.261 |  |
| 4 | RMSE_IDA | 1.940 | 0.577 | 0.565 | 1.940 |  |
| 5 | RMSE_IDA | 3.312 | 3.296 | 3.291 | 3.312 | * |
| 6 | RMSE_IDA | 0.763 | 0.625 | 0.594 | 0.763 |  |
| 7 | RMSE_IDA | 0.751 | 0.678 | 0.659 | 0.752 |  |
| 8 | RMSE_IDA | 1.123 | 1.047 | 1.026 | 1.122 |  |
| 9 | RMSE_IDA | 1.281 | 0.625 | 0.484 | 1.281 |  |
| 10 | RMSE_IDA | 5.752 | 4.766 | 1.961 | 5.753 | * |
| 11 | RMSE_IDA | 1.351 | 1.230 | 1.063 | 1.351 |  |
| 12 | RMSE_IDA | 1.203 | 0.591 | 0.540 | 1.203 |  |
| 13 | RMSE_IDA | 1.007 | 0.919 | 0.686 | 1.007 |  |
| 14 | RMSE_IDA | 0.909 | 0.768 | 0.692 | 0.909 |  |
| 15 | RMSE_IDA | 1.290 | 1.001 | 0.987 | 1.290 |  |
| 16 | RMSE_IDA | 1.283 | 0.692 | 0.650 | 1.283 |  |
| 17 | RMSE_IDA | 0.692 | 0.612 | 0.524 | 0.696 |  |
| 18 | RMSE_IDA | 0.705 | 0.670 | 0.643 | 0.702 |  |
| 19 | RMSE_IDA | 2.986 | 1.072 | 1.025 | 2.986 |  |
| 20 | RMSE_IDA | 0.685 | 0.649 | 0.640 | 0.685 |  |
| * indicates the status of being a outlier. Parameter Setting: SNPs700/Genes70/OverallSamples100000/GtexSamples800/GraphDensity0.05/Weight0.1 | | | | | | |

Here, in replicate1, the performance improves dramatically after p-value adjustment. But in replicate5 and 10, the correlation and RMSE between trueIDA and estimateIDA don’t increase much. In this case, it shows outlier in the boxplot view. More details about the specific replicates are provided in the ‘OutlierExample.xlsx’ file.

Table S4 Combination of self-reported disease and ICD-10 coded diagnosis used for the definition of binary traits

| Trait | Self-reported | | ICD-10 Coding | |
| --- | --- | --- | --- | --- |
| Trait | Coding | Meaning | Coding | Meaning |
| Atrial fibrillation | --- | --- | I48 | Atrial fibrillation |
| Anxiety | 1287 | anxiety/panic attacks | F41 | Other anxiety disorder |
| Breast cancer | 1002 | breast cancer | C50,D05 | Breast cancer, In situ breast cancer |
| CAD | --- | --- | I20-I25 | Ischemic heart disease |
| Depression | 1286 | depression | F32-F33 | Depression |
| Heart failure | 1076 | Heart failure/pulmonary odema | I50 | Heart failure |
| Hypertension | 1065 | hypertension | I10 | Primary hypertension |
| Prostate cancer | 1044 | --- | --- | prostate cancer |
| Stroke | 1081,1082,1083,1086,1583 | stroke, subdural haemorrhage/haematoma, subarachnoid haemorrhage, ischaemic stroke, transient ischaemic attack (tia) | I60-I63 | Stroke |

Table S5 Summary of identified causal graphs for studied traits from exploratory analysis

Table S6 Proportion of small effect genes under different definitions

Table S7 Summary of identified causal graphs for studied traits from selective analysis

Table S8 Stability selection results of inferred gene-gene networks for studied traits with selected gene set

Table S9 Open targets enrichment analysis results under 4 association scores

Table S10 Comparison between our proposed method and the univariate test in identifying potential targets under 4 association scores

Table S11 Novel genes identified from our proposed method compared with the univariate test

Table S12 Comparison of survival analysis on gene sets identified from our method and the univariate test based on the cox proportional-hazard model

| Tissue | Our method | | Univariate test | |
| --- | --- | --- | --- | --- |
|  | Genes | P | Genes | P |
| Breast Mammary | ATXN3 | 6.40E-09 | LRRC37A2 | 3.10E-12 |
|  | CPNE3 | 4.10E-07 | LRRC37A4P | 1.00E+00 |
|  | PTK6 | 2.00E-04 | RPS23 | 1.00E-04 |
|  | GPLD1 | 7.60E-05 | XPNPEP3 | 9.10E-03 |
|  | PRPH2 | 8.20E-08 | GPLD1 | 7.60E-05 |
|  | FAM227B | 8.58E-01 | ATG10 | 1.10E-05 |
|  | XPNPEP3 | 9.10E-03 | ATP6AP1L | 2.20E-03 |
| Whole Blood | GDI2 | 3.00E-04 | NOTCH2 | 4.30E-03 |
|  | DIDO1 | 1.50E-06 | RPS23 | 1.00E-04 |
|  | ANGPTL6 | 1.14E-02 | LRRC37A2 | 3.10E-12 |
|  | NOTCH2 | 4.30E-03 | FLJ45049 | 3.00E-04 |
|  | ZBTB38 | 3.00E-04 | C20orf158 | 1.50E-06 |
|  | RPS23 | 1.00E-04 | MAPK8IP1 | 8.00E-03 |
|  | SLC22A5 | 2.60E-13 | LRDD | 6.20E-06 |
|  | LRDD | 6.20E-06 | LRRC37A | 6.00E-04 |

For LRRC37A4P, no available data from the reference database, so we set the p value to 1 here.

Table S13 Summary of inferred causal graphs for multiple traits analysis

Table S14 Pathway enrichment analysis of identified core genes for studied traits

Table S15 Drug enrichment analysis results for studied binary traits based on identified core genes

Table S16 Transcription factor enrichment analysis results for studied binary traits based on identified core genes

Table S17 Number of causally relevant genes of studied phenotypes in different tissues after incorporating external knowledge from SIGNOR

### Figures

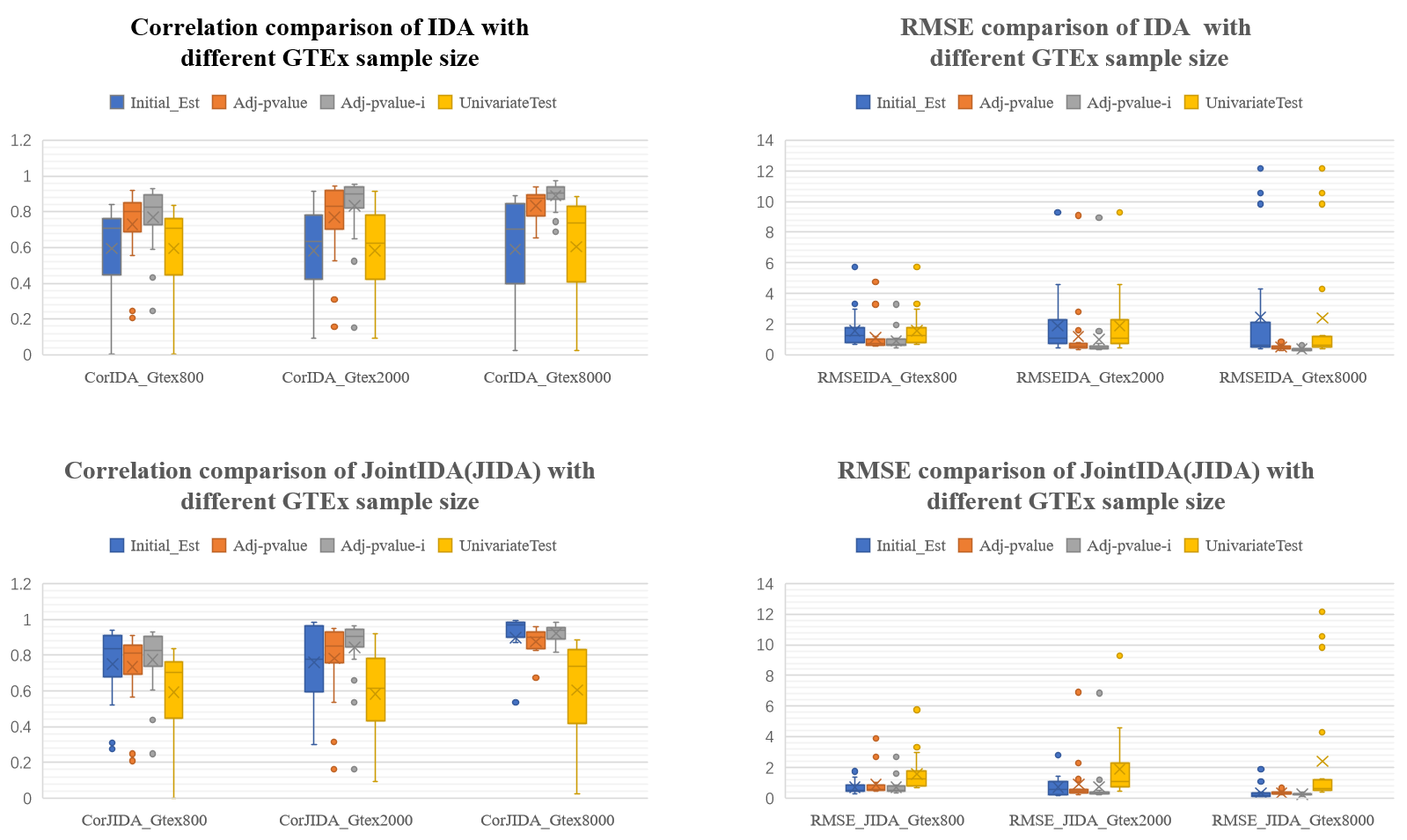

Fig.S1 The Correlation and root mean square error (RMSE) between true value and estimated value, for different GTEx sample size.

Parameter Setting: SNPs700/Genes70/OverallSamples100000/GraphDensity0.05/**MinWeight0.1**

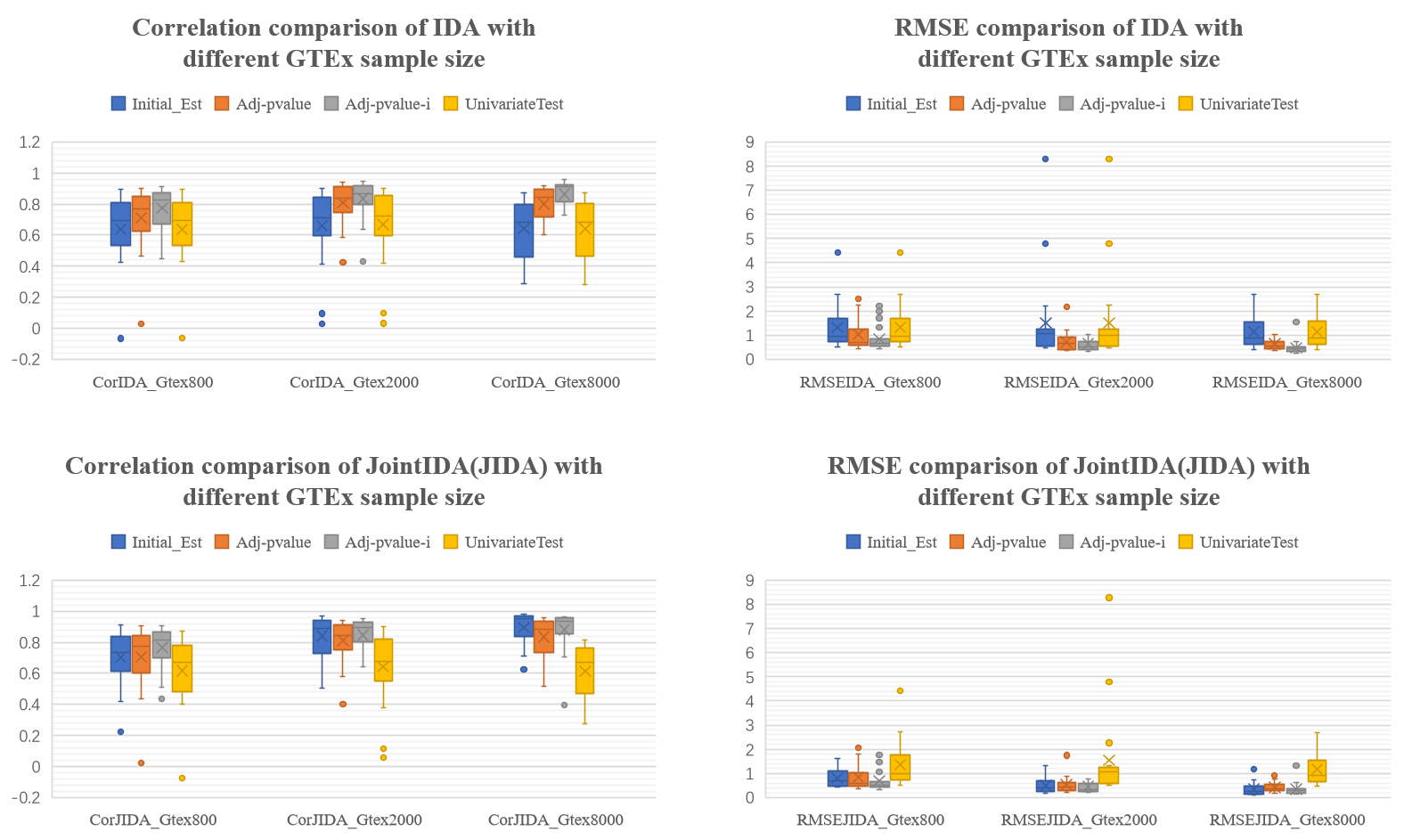

Fig.S2 The Correlation and root mean square error (RMSE) between true value and estimated value, for different GTEx number.

Parameter Setting: SNPs700/Genes70/OverallSamples100000/GraphDensity0.05/**MinWeight0.3**

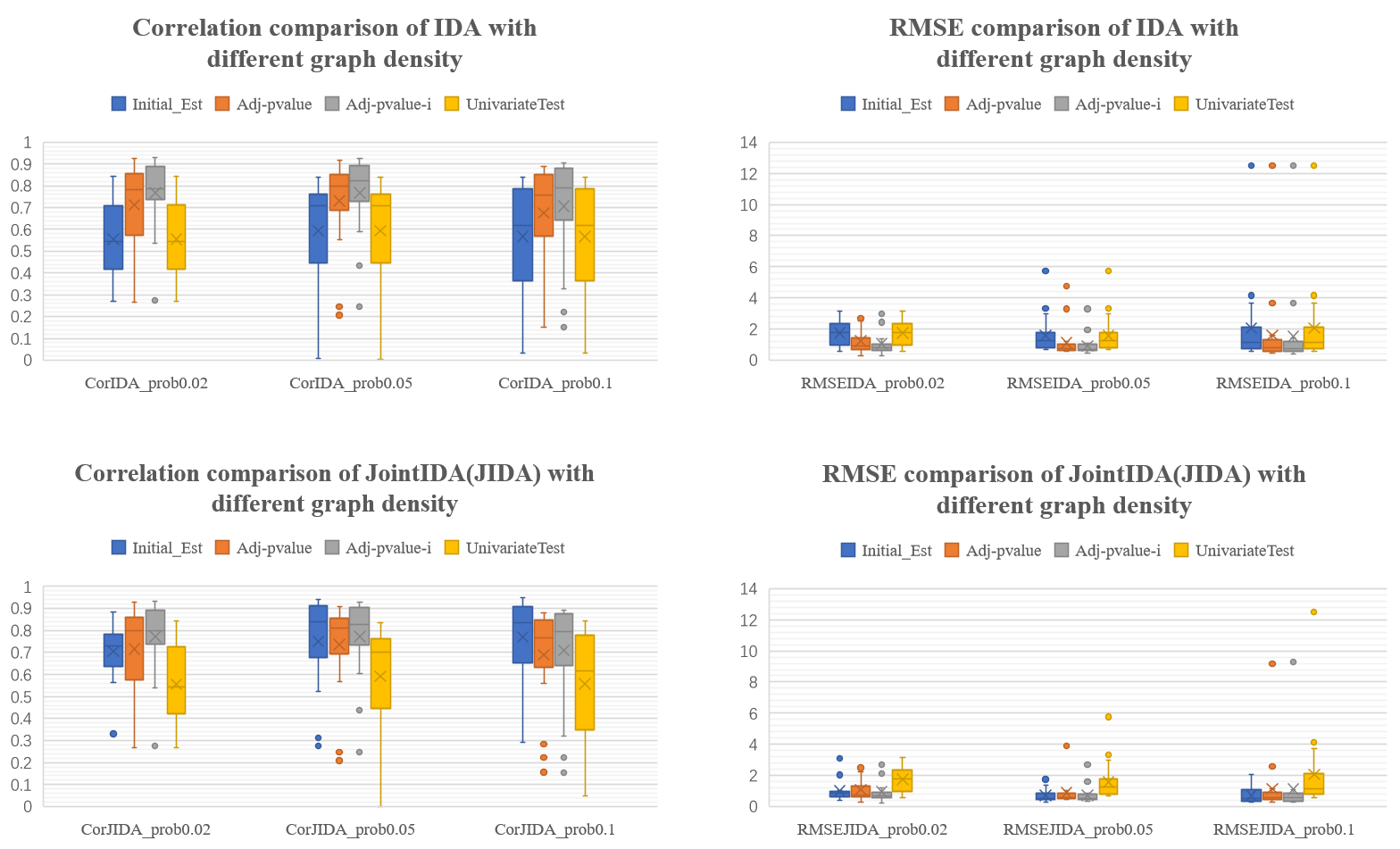

Fig.S3 The Correlation and root mean square error (RMSE) between true value and estimated value, for different graph density.

Parameter Setting: SNPs700/Genes70/OverallSamples100000/GTExSamples800/ /**MinWeight0.1**

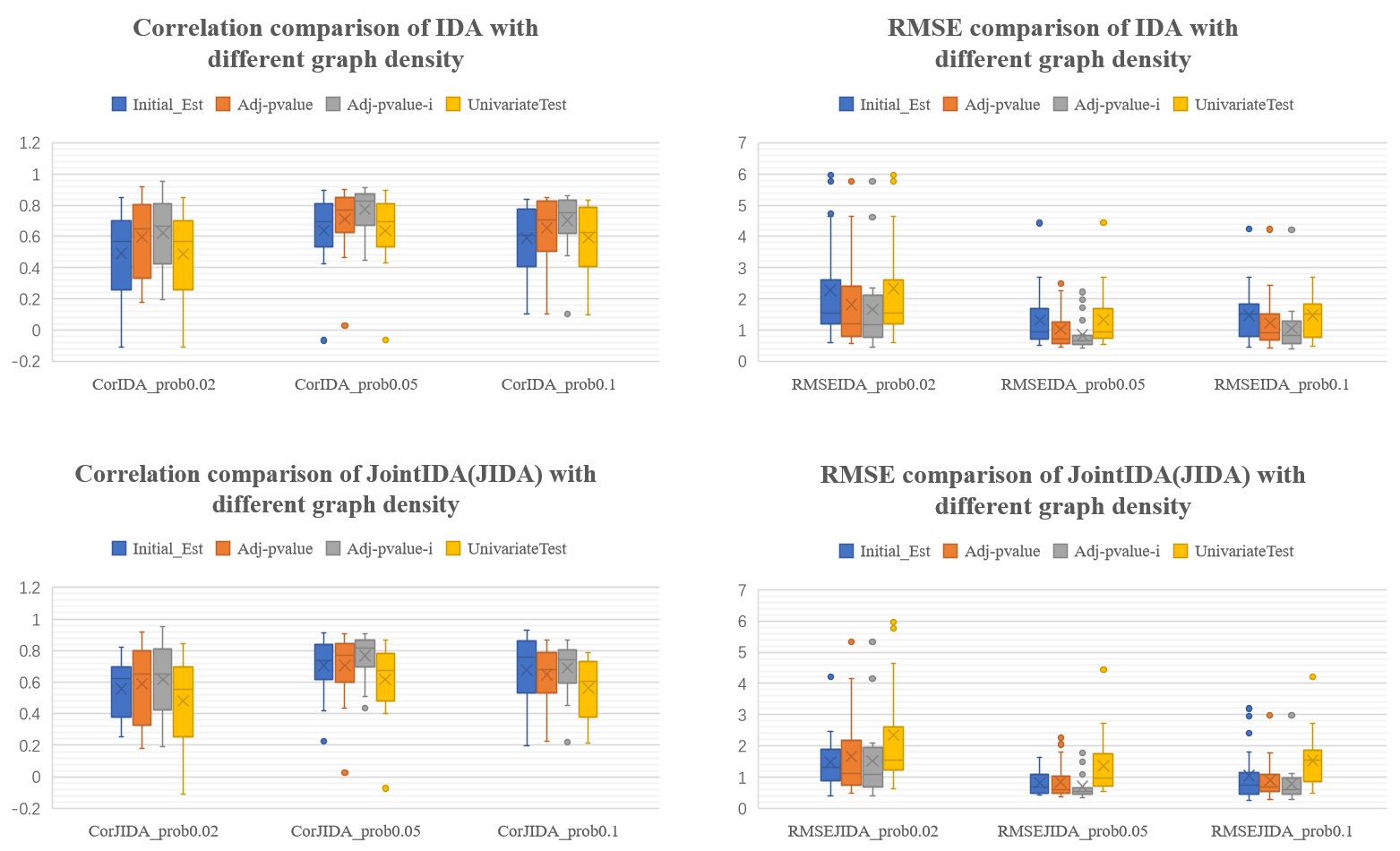

Fig.S4 The Correlation and root mean square error (RMSE) between true value and estimated value, for different graph density.

Parameter Setting: SNPs700/Genes70/OverallSamples100000/GTExSamples800/ /**MinWeight0.3**

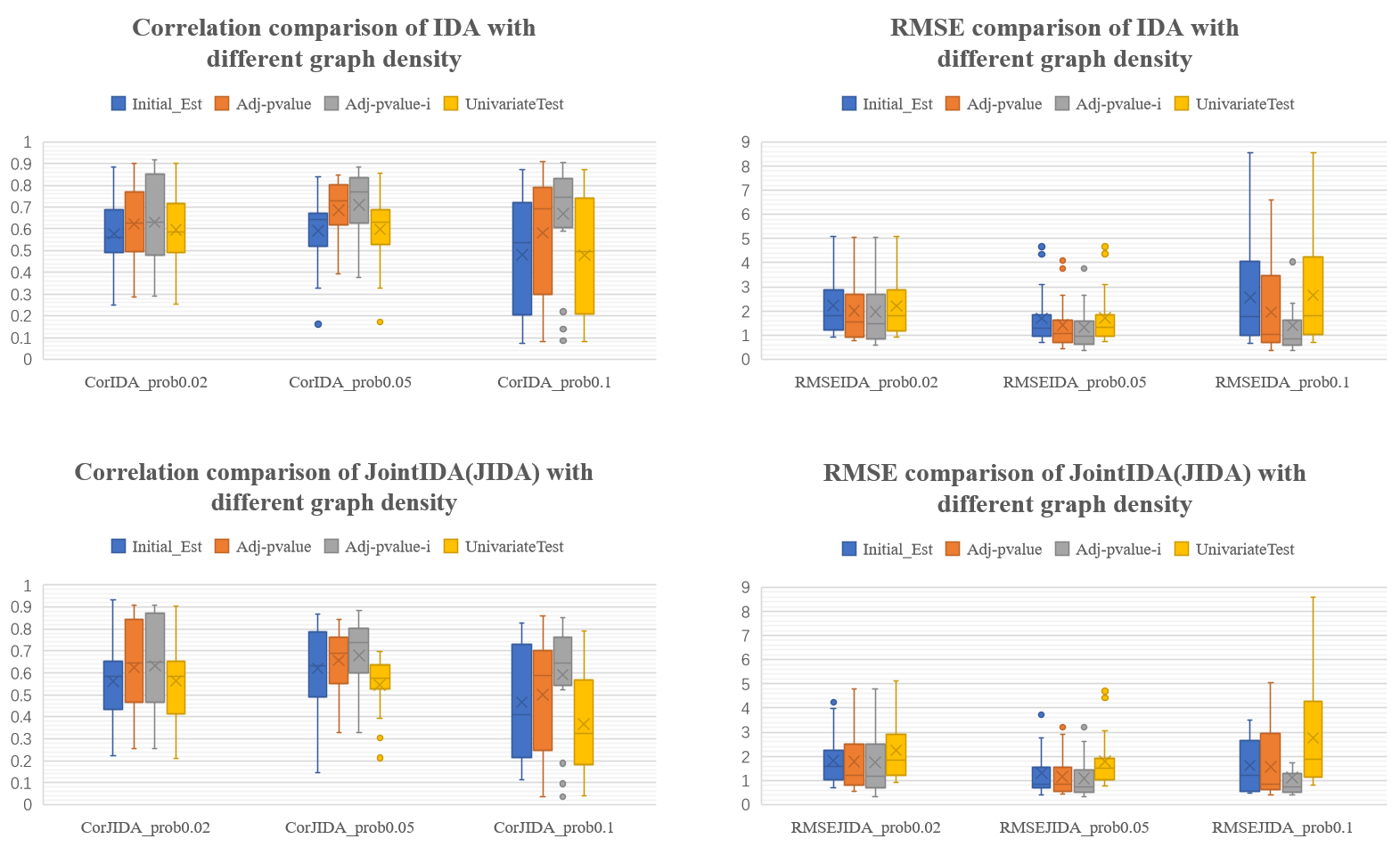

Fig.S5 The Correlation and root mean square error (RMSE) between true value and estimated value, for different graph density.

Parameter Setting: SNPs700/Genes70/OverallSamples100000/GTExSamples800/ /**MinWeight0.5**

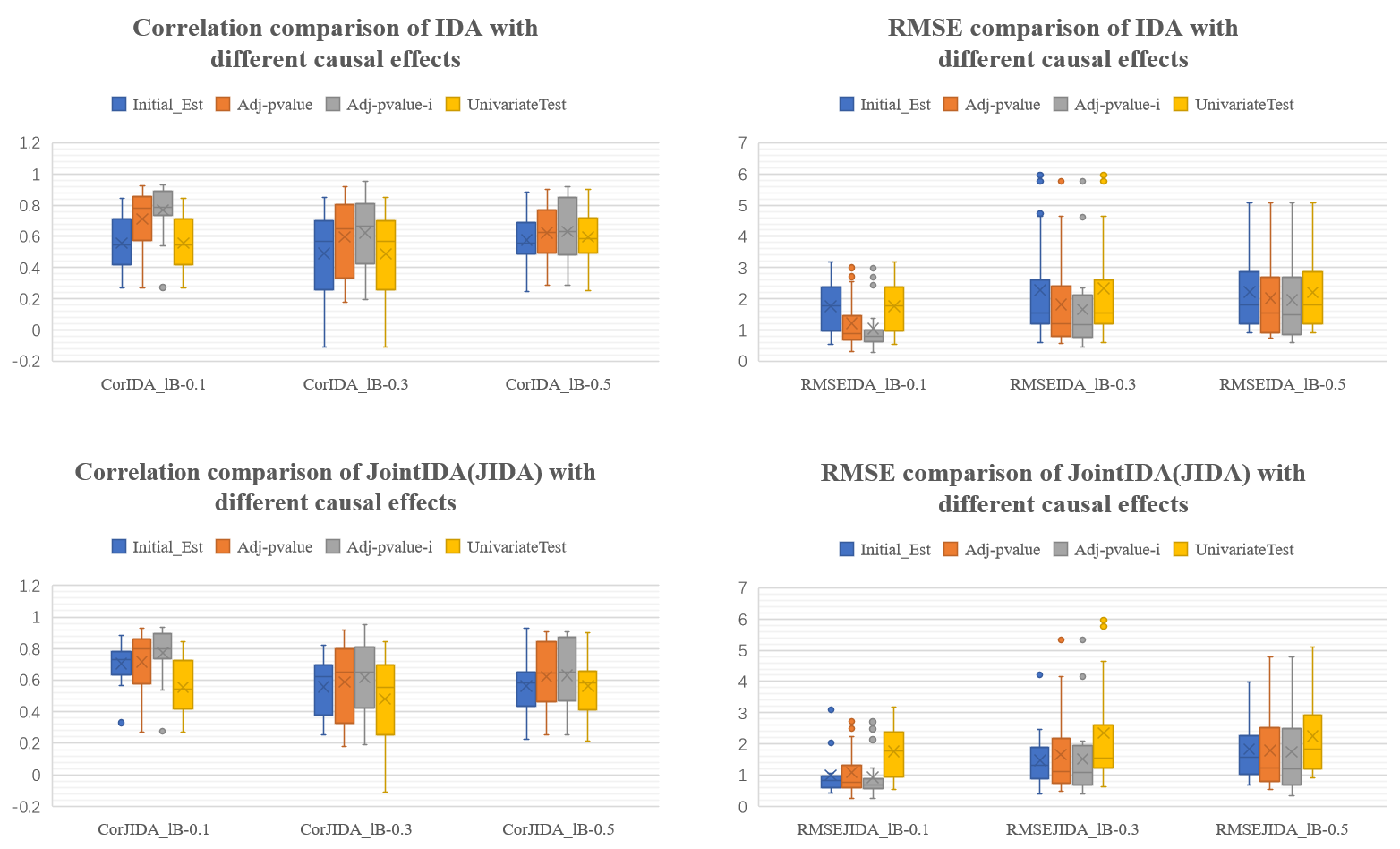

Fig.S6 The Correlation and root mean square error (RMSE) between true value and estimated value, for different causal effects.

Parameter Setting: SNPs700/Genes70/OverallSamples100000/GTExSamples800/**GraphDensity0.02**

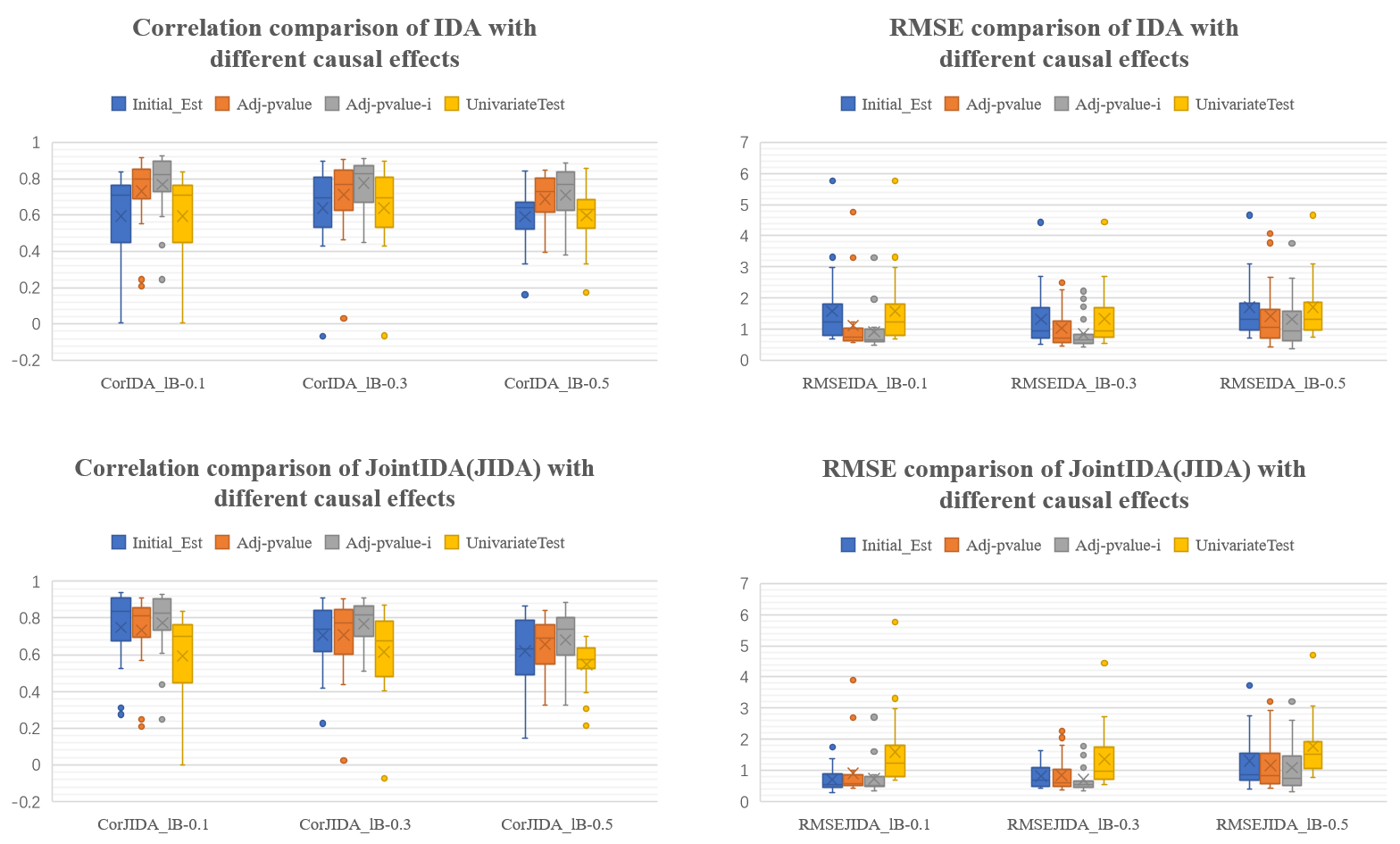

Fig.S7 The Correlation and root mean square error (RMSE) between true value and estimated value, for different causal effects.

Parameter Setting: SNPs700/Genes70/OverallSamples100000/GTExSamples800/**GraphDensity0.05**

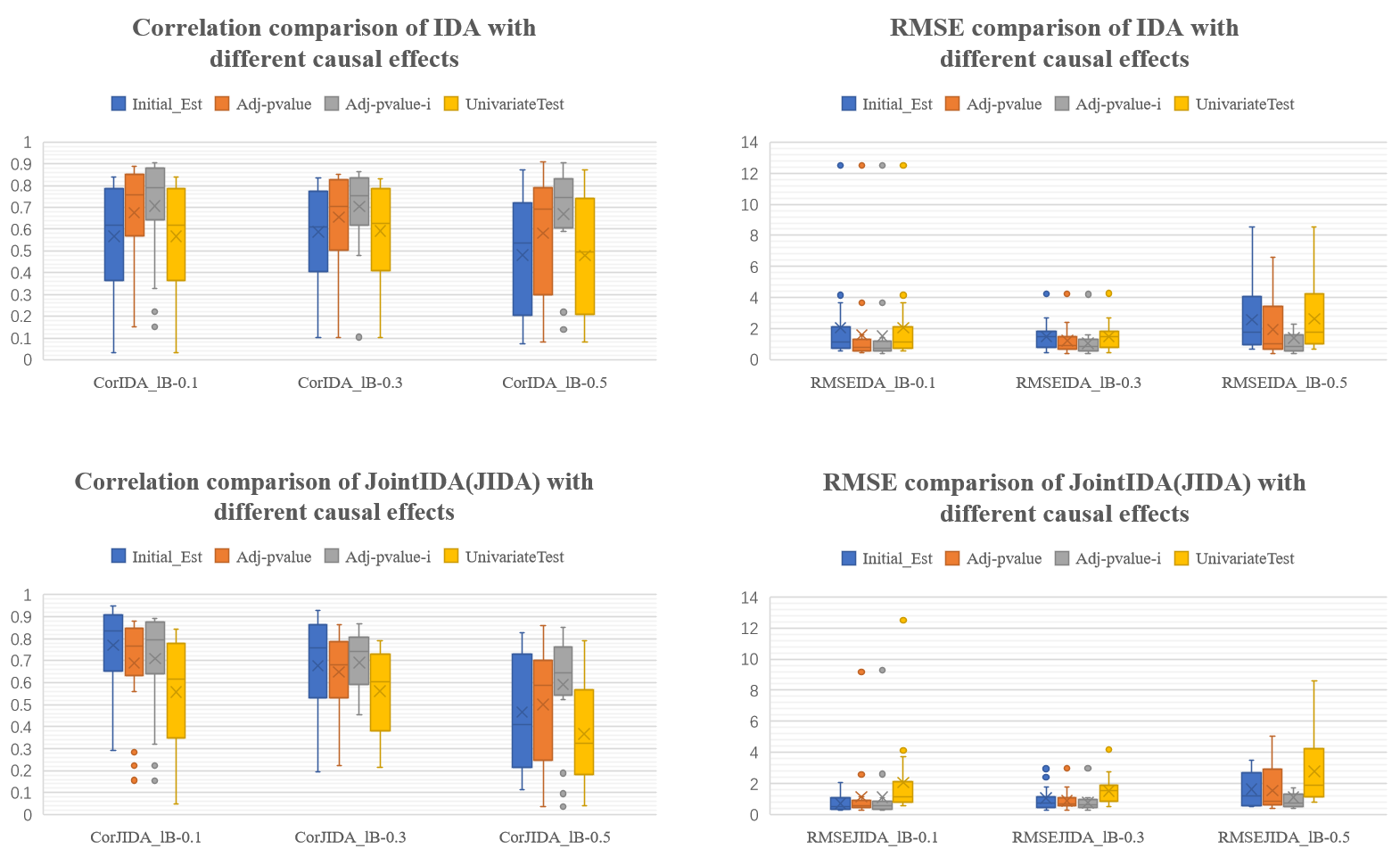

Fig.S8 The Correlation and root mean square error (RMSE) between true value and estimated value, for different causal effects.

Parameter Setting: SNPs700/Genes70/OverallSamples100000/GTExSamples800/**GraphDensity0.1**

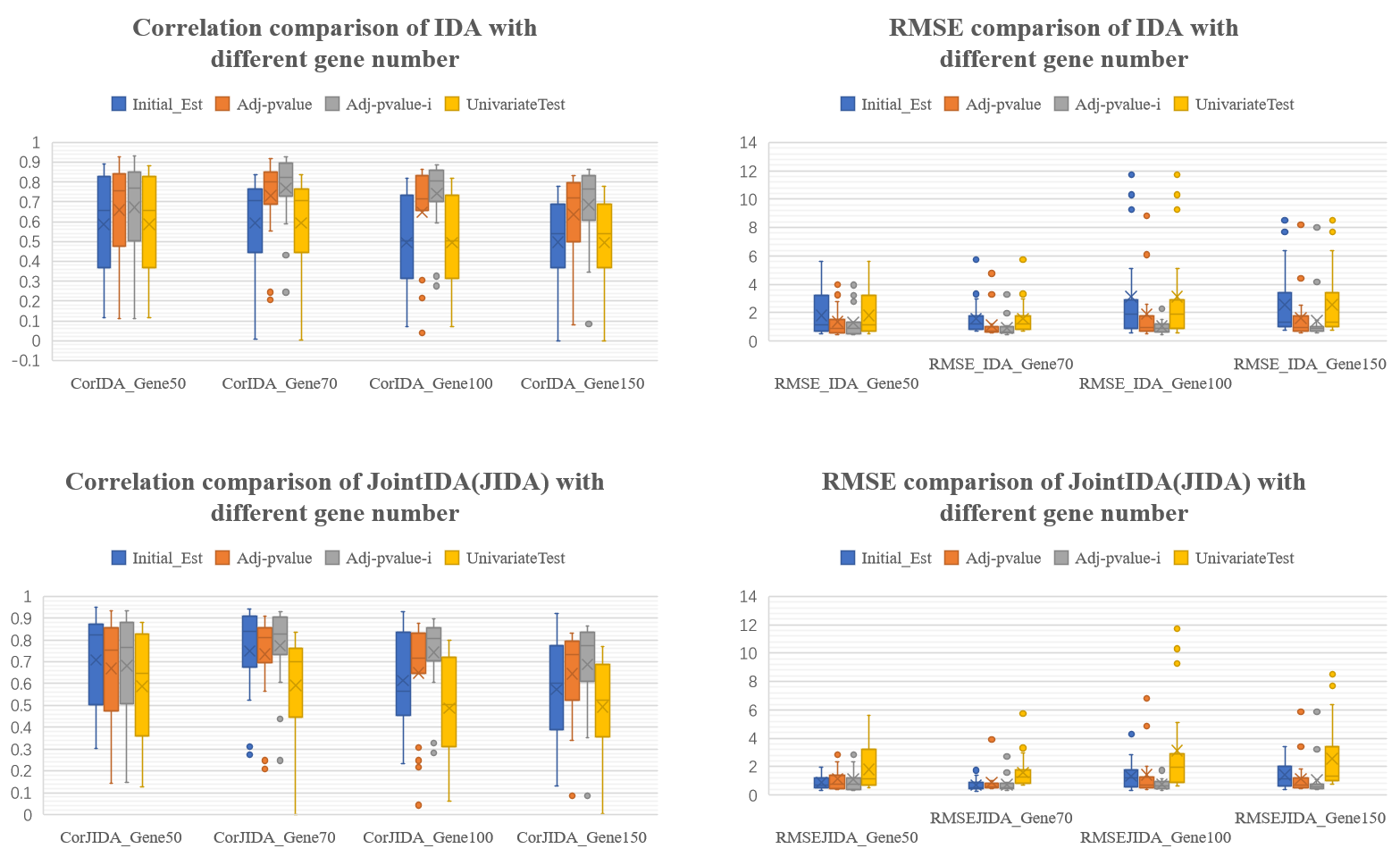

Fig.S9 The Correlation and root mean square error (RMSE) between true value and estimated value, for different gene number.

Parameter Setting: OverallSamples100000/GTExSamples800/GraphDensity0.05/**MinWeight0.1**

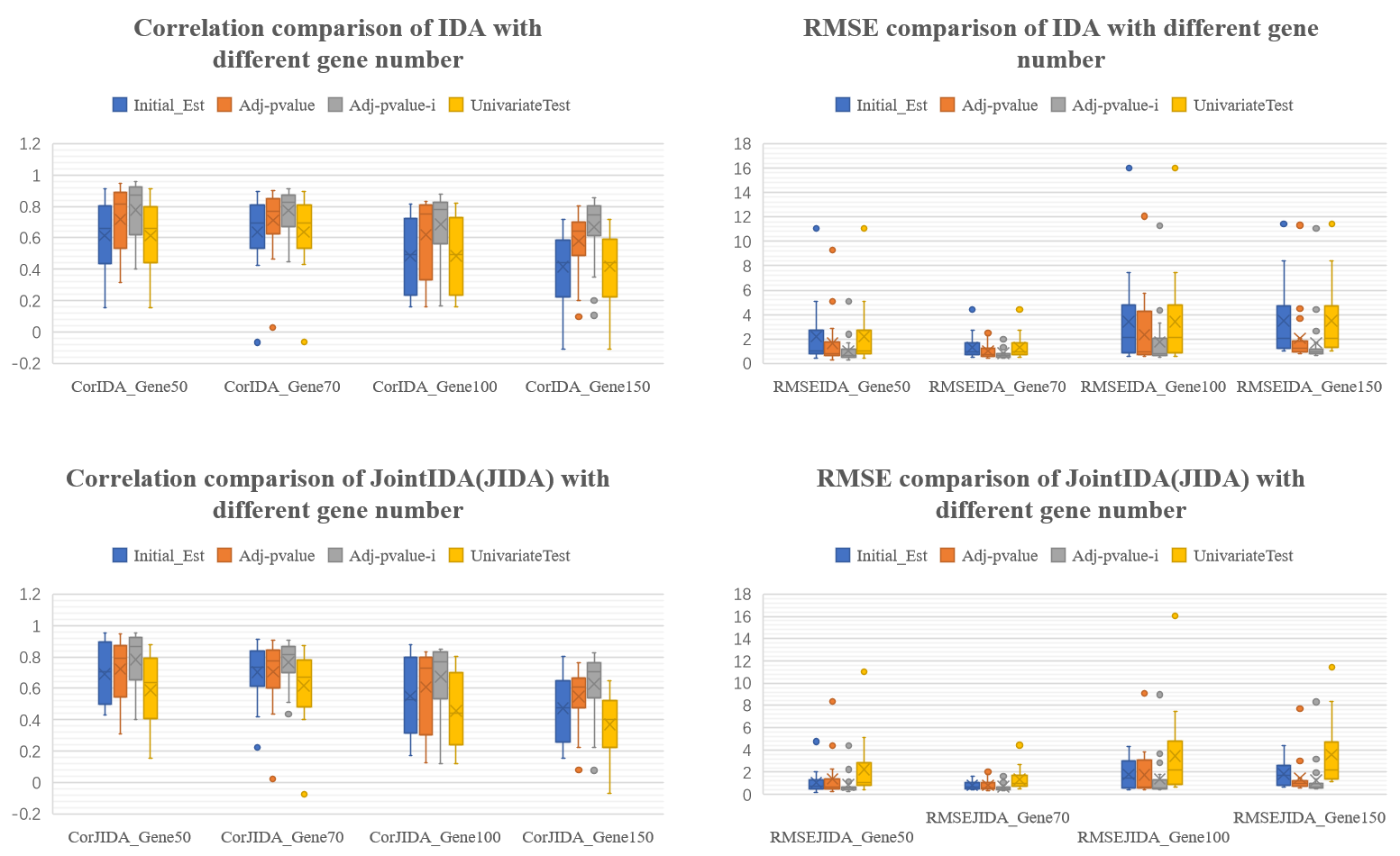

Fig.S10 The Correlation and root mean square error (RMSE) between true value and estimated value, for different gene number.

Parameter Setting: OverallSamples100000/GTExSamples800/GraphDensity0.05**/MinWeight0.3**

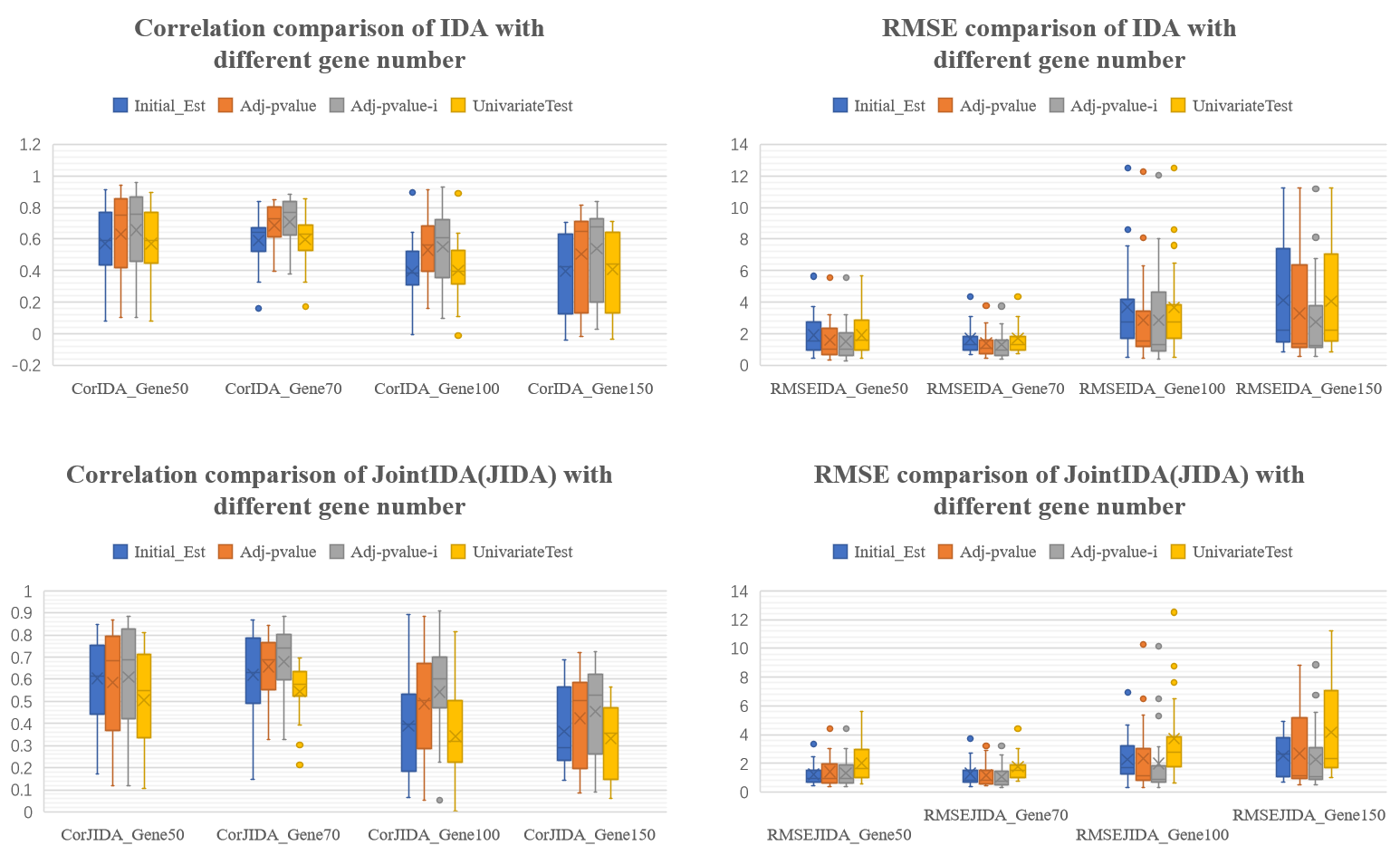

Fig.S11 The Correlation and root mean square error (RMSE) between true value and estimated value, for different gene number.

Parameter Setting: OverallSamples100000/GTExSamples800/GraphDensity0.05/**MinWeight0.5**

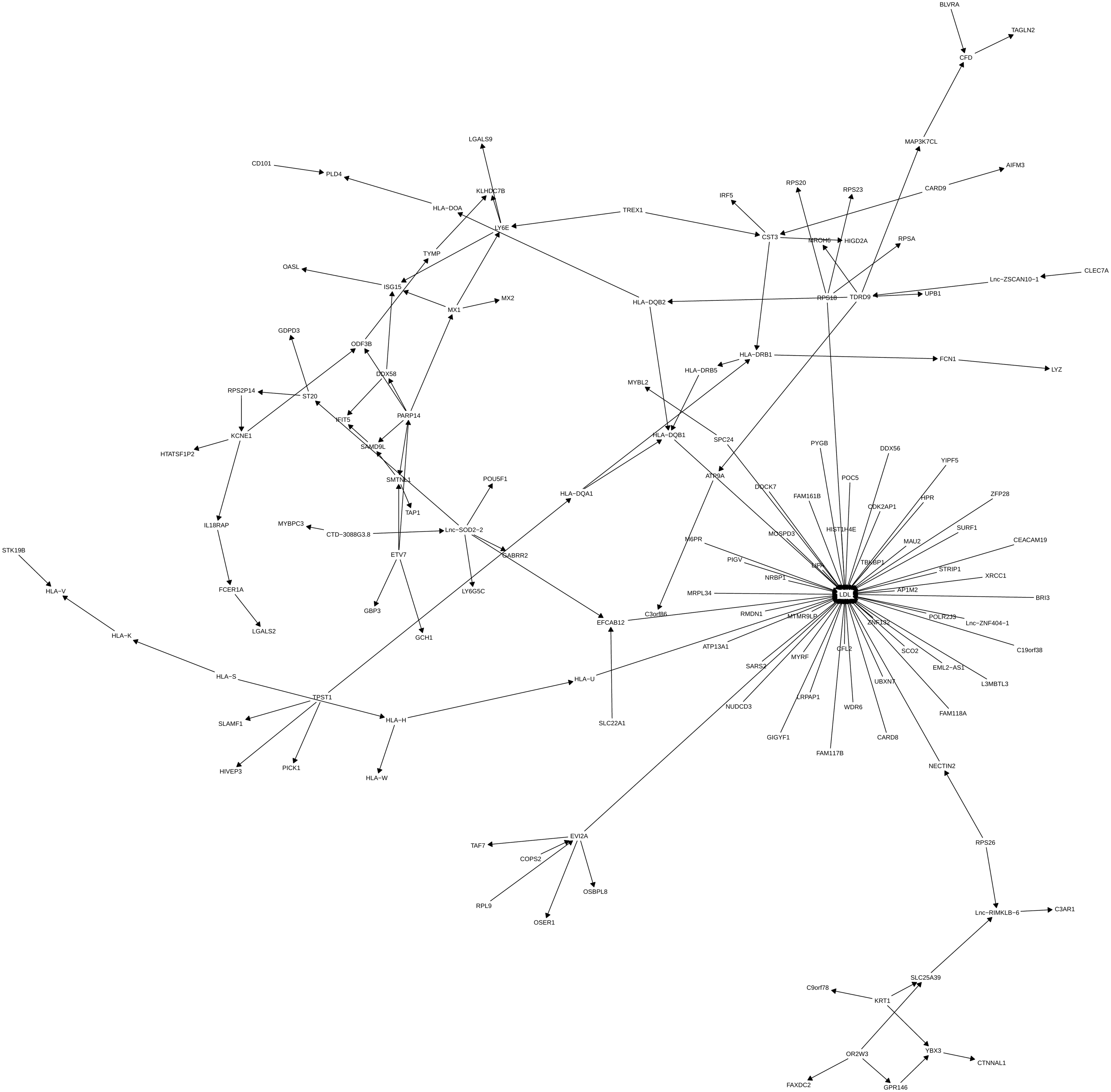

Fig S12 The estimated causal graph for LDL (low density lipoprotein) in the whole blood
